# An international factorial vignette-based survey of intubation decisions in acute hypoxemic respiratory failure

**DOI:** 10.1101/2024.04.16.24305906

**Authors:** Christopher J Yarnell, Arviy Paranthaman, Peter Reardon, Federico Angriman, Thiago Bassi, Giacomo Bellani, Laurent Brochard, Harm Jan De Grooth, Laura Dragoi, Syafruddin Gaus, Paul Glover, Ewan C Goligher, Kimberley Lewis, Baoli Li, Hashim Kareemi, Bharath Kumar Tirupakuzhi Vijayaraghavan, Sangeeta Mehta, Ricard Mellado-Artigas, Julie Moore, Idunn Morris, Georgiana Roman-Sarita, Tai Pham, Jariya Sereeyotin, George Tomlinson, Hannah Wozniak, Takeshi Yoshida, Rob Fowler, Canadian Critical Care Trials Group

**Affiliations:** Department of Critical Care Medicine, Scarborough Health Network, Toronto, Ontario, Canada; Scarborough Health Network Research Institute, Toronto, Ontario, Canada; Keenan Research Centre for Biomedical Research, Li Ka Shing Knowledge Institute, St Michael’s Hospital, Unity Health Toronto, Canada; Interdepartmental Division of Critical Care Medicine, University of Toronto, Toronto, Canada; Department of Medicine, Division of Respirology, University Health Network; Department of Medicine, Sinai Health System, Toronto, Canada; Department of Medicine, University of Toronto, Toronto, Canada; Department of Respiratory Therapy, University Health Network, Toronto, Canada; Department of Medicine, University Health Network, Toronto, Canada; Institute for Clinical Evaluative Sciences, Toronto, Canada; Institute of Health Policy, Management and Evaluation, University of Toronto, Canada; Sunnybrook Health Sciences Centre, Toronto, Canada; Division of Critical Care, Department of Medicine, McMaster University, Hamilton, Canada; Department of Health Research, Methods, Evidence, and Impact, McMaster University, Hamilton, Canada; Service de Medecine Intensive-Réanimation, Assistance Publique–Hôpitaux de Paris, Hôpital de Bicêtre, DMU CORREVE, FHU SEPSIS, Groupe de Recherche CARMAS, Le Kremlin-Bicêtre, France; Universite Paris-Saclay, Université de Versailles Saint-Quentin-en-Yvelines, Université Paris-Sud, Inserm U1018, Equipe d’Epidemiologie Respiratoire Intégrative, Centre d’ Epidémiologie et de Santé des Populations, Villejuif, France; Surgical ICU, Department of Anesthesiology, Hospital Clínic de Barcelona, Barcelona (Spain); Lawrence S. Bloomberg Faculty of Nursing, University of Toronto, Toronto, Canada; Trent-Fleming School of Nursing, Trent University, Peterborough, Ontario; Department of Nursing, Mount Sinai Hospital, Toronto, Canada; Nepean Clinical School, University of Sydney, New South Wales, Australia; Department of Intensive Care Medicine, Nepean Hospital, Sydney, Australia; Department of Physiology, University of Toronto, Toronto, Canada; Intensive Care Center, UMC Utrecht, Utrecht, The Netherlands; Department of Anesthesiology, Pharmacology, Intensive Care and Emergency Medicine, Geneva University Hospitals, Geneva, Switzerland; Centre for Medical Sciences (CISMed), University of Trento, 38122, Trento, Italy; Department of Critical Care Medicine, Apollo Hospitals, Chennai, India and Centre for Inflammation Research, University of Edinburgh, U.K.; Anaesthesia and Intensive Care, Santa Chiara Hospital, APSS, Trento, Italy; Department of Anesthesiology, Division of Critical Care Medicine, King Chulalongkorn Memorial Hospital and Faculty of Medicine, Chulalongkorn University, Bangkok, Thailand; Department of Emergency Medicine, University of British Columbia, Vancouver, Canada; The Department of Anesthesiology and Intensive Care Medicine, Osaka University Graduate School of Medicine, Suita, Japan; Department of Anesthesiology, Intensive Care, and Pain Management; Faculty of Medicine, Hasanuddin University, Makassar, Indonesia; Northern Ontario School of Medicine, Timmins and District Hospital, Timmins, Ontario, Canada

**Keywords:** Hypoxemic Respiratory Failure, Endotracheal Intubation, Healthcare Surveys, Multilevel Analyses

## Abstract

**Purpose:** Intubation is a common procedure in acute hypoxemic respiratory failure (AHRF), with minimal evidence to guide decision-making. We conducted a survey of when to intubate patients with AHRF to measure the influence of clinical variables on intubation decision-making and quantify variability.

**Methods:** We developed an anonymous factorial vignette-based web survey to ask clinicians involved in the decision to intubate “Would you recommend intubation?” Respondents selected an ordinal recommendation from a 5-point scale ranging from “Definite no” to “Definite yes” for up to 10 randomly allocated vignettes. We disseminated the survey through clinical and academic societies, analyzed responses using Bayesian proportional odds modeling with clustering by individual, country, and region, and reported mean odds ratios (OR) with 95% credible intervals (CrI).

**Results:** Between September 2023 and January 2024, 2,294 respondents entered 17,235 vignette responses in 74 countries [most common: Canada (29%), USA (26%), France (9%), Japan (8%), and Thailand (5%)]. Respondents were attending physicians (63%), nurses (13%), trainee physicians (9%), respiratory therapists (9%), other (6%). Lower oxygen saturation, higher inspired oxygen fraction, non-invasive ventilation compared to high-flow, tachypnea, neck muscle use, abdominal paradox, drowsiness, and inability to obey were associated with increased odds of intubation; diagnosis, vasopressors, and duration of symptoms were not. Within a country the odds of recommending intubation changed between clinicians by an average factor of 2.60, while changing between countries within a region changed it by 1.56.

**Conclusion:** In this international, interprofessional survey of more than 2000 practicing clinicians, intubation for patients with AHRF was mostly decided based on oxygenation, breathing pattern, and consciousness, but there was important variation across individuals and countries.

## Introduction

When to intubate people with acute hypoxemic respiratory failure (AHRF) is an important and common dilemma for clinicians, but optimal criteria to guide initiation are unknown(1). Uncertainty in this decision is important, because earlier intubation may unnecessarily expose patients to risks of periprocedural shock and cardiac arrest(2), laryngeal and dental trauma(3), infection(4), ventilator-induced lung injury(5), immobility and weakness(6), and delirium and post-traumatic stress disorder(7); later intubation may increase the risk of death due to physiologically dangerous intubating conditions(8), worsening of self-inflicted lung injury(9), or decompensated respiratory failure progressing to cardiac arrest(10).

In qualitative research and surveys, clinicians have highlighted the importance of physiologic variables such as degree of hypoxemia and breathing pattern in deciding when to intubate(11–13). However, there is variation in the use of invasive ventilation by location, time period, and patient race(14–18). No research yet investigates the relationships between physiological variables and intubation decisions, and the extent of variability in decision-making between individuals and around the world is unknown.

To address these uncertainties, we conducted an international, scenario-based electronic survey to quantify variability in practice and measure the influence of clinical variables on intubation decisions for patients with AHRF.

## Methods

We designed and disseminated an anonymous electronic survey using established methods.(19,20) We obtained approval from the Scarborough Health Network research ethics board (#ICU-23-009). Completion of the survey implied consent. We pre-registered the analysis plan prior to completion of data collection(21) and analytic code is publicly available.(cite doi)

### Participants

We targeted a global population of clinicians who self-identified as “involved in the decision to initiate invasive ventilation for respiratory failure as part of their clinical practice.” Because we targeted a global population of diverse professions, we could not ascertain a denominator for calculating response rates.(22)

### Development and piloting

We identified key physiologic variables hypothesized to influence intubation decisions using randomized trials(23), observational studies of thresholds for intubation (15,24), research on mechanical ventilation weaning physiology(25–28), and our clinical experience with patients experiencing respiratory failure. We performed clinical sensibility testing with 8 clinicians who are involved in intubation decisions (2 respiratory therapists, 4 physicians, 2 nurses) and revised the survey based on that feedback.(19,29)

### Survey design

We designed a factorial vignette-based survey.(30–32) Variables were patient age, premorbid frailty(33), diagnosis, oxygen saturation, inspired oxygen fraction, oxygen device, respiratory rate, breathing pattern, norepinephrine use, level of consciousness, and duration in current state(eTable 1). Variables were randomly combined to generate vignettes for respondents. This design guarded against question order bias(34), allowed for detailed exploration of interactions(35), and minimized the impact of bias due to the perception of being either observed (Hawthorne effect) or evaluated (sentinel effect)(30). After discretizing continuous variables (eg. respiratory rate) and introducing clinically relevant covariance (eg. removing norepinephrine from vignettes with a diagnosis of COVID), there were 1.25 million potential vignettes.

The survey began with a questionnaire to gather respondent demographics (eFigures 1 and 2) including clinical role, primary clinical area of practice, duration in practice, and country of practice. We then presented respondents with 10 randomly allocated tabular vignettes (eFigure 3).

For each vignette, respondents were asked “Would you recommend intubation and invasive ventilation?” and selected an ordinal response from “Definite no”, “Probable no”, “Uncertain”, “Probable yes”, to “Definite yes.” Respondents could also indicate if any of 6 additional information types would be helpful for their decision (arterial blood gas, chest radiography, positive end-expiratory pressure (PEEP), tidal volume measurement, esophageal manometry, or more observation time, eFigure 4).

### Translation and dissemination

The English survey was translated by fluent clinicians into Indonesian, Chinese (simplified), French, German, Italian, Japanese, Portuguese, Spanish, and Thai. The survey was disseminated through and/or endorsed by several societies and networks listed in the Acknowledgments.

### Descriptive analyses

We reported the number of respondents according to characteristics. Regions, sub-regions, and countries were defined using the ISO 3166 country codes.(36) We reported the total, number per respondent, the distribution of responses by respondent characteristics, and the number requesting each additional information type.

### Primary analysis

We analyzed the ordinal intubation recommendation outcome with a Bayesian multilevel proportional odds model(37,38). The model clustered observations by region, country, and individual to account for repeated measurements. Multilevel structure also allowed for country-level odds ratios to be related to the number of respondents, by shrinking odds ratios for countries with few respondents towards 1. This is a quantitative improvement on excluding sparse categories or combining them into an “Other” category(39).

### Data processing and predictors

We categorized continuous variables to allow non-linearity with interpretable coefficients. We used all respondent and vignette data as predictors, including bivariate interactions between all vignette variables. Introducing interactions increased the number of parameters from 47 to 501, therefore we chose prior distributions that prevent overfitting(40) (see Supplement).

### Outputs

From the primary analysis, we reported odds ratios for main effects, interactions, and combinations of country- and region-level random intercepts. We quantified variability between individuals, countries, and regions using the median odds ratio.(41,42)

### Secondary analyses

#### Requests for additional information

We performed Bayesian multilevel logistic regression, with the same structure as the primary analysis, for whether each type of additional information was requested.

#### Comparing with observed data

We compared survey responses to observed data to address the common criticism of vignette-based surveys that their responses do not reflect real-world decision-making. To do so, we constructed a cohort of all observations of patients receiving oxygen via high-flow nasal cannula or non-invasive ventilation from the Medical Information Mart for Intensive Care IV (MIMIC-IV) database, similar to a prior study(15). For each observation, we used the model from the primary analysis to predict survey responses. Then, we compared the predicted survey responses to the observed outcome of intubation within 3, 8, or 24 hours.(15,43,44) We reported discrimination and calibration of the predicted recommendations versus observed intubation rates (Supplement, section 10).

## Sensitivity analyses

We repeated the primary analysis using only respondents who completed 10 scenarios. We did this because some users were prematurely disconnected during high survey traffic. If they reattempted the survey, their data appeared as two separate respondents. Given that it is unlikely that someone who completed 10 scenarios would reattempt the survey, the 10-response subset likely consists of unique respondents.

We also repeated the primary analysis for responses restricted to each scenario diagnosis. This amounted to forcing an interaction between diagnosis and every other variable in the model. In a final sensitivity analysis, we assessed the proportional odds assumption.

## Computational details and reporting

The survey interface was coded in R and shiny(31,45), and stored responses in REDcap.(46,47) We used R and the brms package for all Bayesian analyses.(48,49) Bayesian models used 4 chains of 1000 iterations each, split between warm-up and sampling. We summarized posterior distributions with means and 95% credible intervals (CrI), used complete case analysis for modeling, and defined the region of practical equivalence as an odds ratio of 0.9 to 1.1.(50)

## Results

### Respondents

Between September 27, 2023 and January 11, 2024, 2,294 clinicians from 74 countries involved in the decision to intubate responded to the survey (Table 1). The top 5 countries were Canada (659, 29%), United States (597, 26%), France (199, 9%), Japan (187, 8%), and Thailand (121, 5%) (eFigure 5). Overall, 711 (31%) completed surveys in a non-English language, most commonly French (258, 11%), Japanese (174, 8%), and Spanish (107, 5%).

**Table 1.**

|  | Total | Northern America | Europe | Asia | Oceania | Latin America and<br>the Caribbean | Africa |
| --- | --- | --- | --- | --- | --- | --- | --- |
| **Total** | 2294 | 1256 | 458 | 421 | 72 | 68 | 19 |
| **Survey language** |  |  |  |  |  |  |  |
| Bahasa Indonesia | < 5 (0%) | 0 (0%) | 0 (0%) | < 5 (0%) | 0 (0%) | 0 (0%) | 0 (0%) |
| Chinese (simplified) | < 5 (0%) | < 5 (0%) | 0 (0%) | < 5 (1%) | 0 (0%) | 0 (0%) | 0 (0%) |
| English | 1583 (69%) | 1222 (97%) | 113 (25%) | 154 (37%) | 72 (100%) | 10 (15%) | 12 (63%) |
| German | 10 (0%) | 0 (0%) | 10 (2%) | 0 (0%) | 0 (0%) | 0 (0%) | 0 (0%) |
| French | 258 (11%) | 31 (2%) | 218 (48%) | 0 (0%) | 0 (0%) | < 5 (3%) | 7 (37%) |
| Italian | 58 (3%) | 0 (0%) | 58 (13%) | 0 (0%) | 0 (0%) | 0 (0%) | 0 (0%) |
| Japanese | 174 (8%) | 0 (0%) | 0 (0%) | 174 (41%) | 0 (0%) | 0 (0%) | 0 (0%) |
| Portuguese | 10 (0%) | 0 (0%) | < 5 (0%) | 0 (0%) | 0 (0%) | 9 (13%) | 0 (0%) |
| Spanish | 107 (5%) | < 5 (0%) | 58 (13%) | 0 (0%) | 0 (0%) | 47 (69%) | 0 (0%) |
| Thai | 88 (4%) | 0 (0%) | 0 (0%) | 88 (21%) | 0 (0%) | 0 (0%) | 0 (0%) |
| **Specialty** |  |  |  |  |  |  |  |
| Critical care medicine | 1825 (80%) | 1051 (84%) | 385 (84%) | 246 (58%) | 67 (93%) | 61 (90%) | 15 (79%) |
| Anesthesia | 302 (13%) | 80 (6%) | 91 (20%) | 116 (28%) | 8 (11%) | < 5 (3%) | 5 (26%) |
| Emergency medicine | 340 (15%) | 192 (15%) | 46 (10%) | 89 (21%) | < 5 (1%) | 7 (10%) | 5 (26%) |
| Other | 202 (9%) | 97 (8%) | 24 (5%) | 73 (17%) | 0 (0%) | 5 (7%) | < 5 (16%) |
| **Role** |  |  |  |  |  |  |  |
| Nurse | 310 (14%) | 214 (17%) | 58 (13%) | 12 (3%) | 24 (33%) | < 5 (1%) | < 5 (5%) |
| Attending physician | 1437 (63%) | 689 (55%) | 331 (72%) | 323 (77%) | 32 (44%) | 49 (72%) | 13 (68%) |
| Other clinical role | 146 (6%) | 106 (8%) | 13 (3%) | 18 (4%) | < 5 (4%) | 6 (9%) | 0 (0%) |
| Trainee physician | 204 (9%) | 89 (7%) | 40 (9%) | 56 (13%) | 11 (15%) | < 5 (6%) | < 5 (21%) |
| Respiratory therapist | 197 (9%) | 158 (13%) | 16 (3%) | 12 (3%) | < 5 (3%) | 8 (12%) | < 5 (5%) |
| **Duration in practice** |  |  |  |  |  |  |  |
| 0-5 years | 662 (29%) | 404 (32%) | 111 (24%) | 109 (26%) | 13 (18%) | 21 (31%) | < 5 (21%) |
| 6-10 years | 440 (19%) | 232 (18%) | 103 (22%) | 76 (18%) | 18 (25%) | < 5 (6%) | 7 (37%) |
| 11-20 years | 486 (21%) | 243 (19%) | 113 (25%) | 99 (24%) | 12 (17%) | 14 (21%) | 5 (26%) |
| 20-60 years | 686 (30%) | 364 (29%) | 127 (28%) | 136 (32%) | 28 (39%) | 29 (43%) | < 5 (11%) |
| Missing | 20 (1%) | 13 (1%) | < 5 (1%) | < 5 (0%) | < 5 (1%) | 0 (0%) | < 5 (5%) |

**Table 2.**
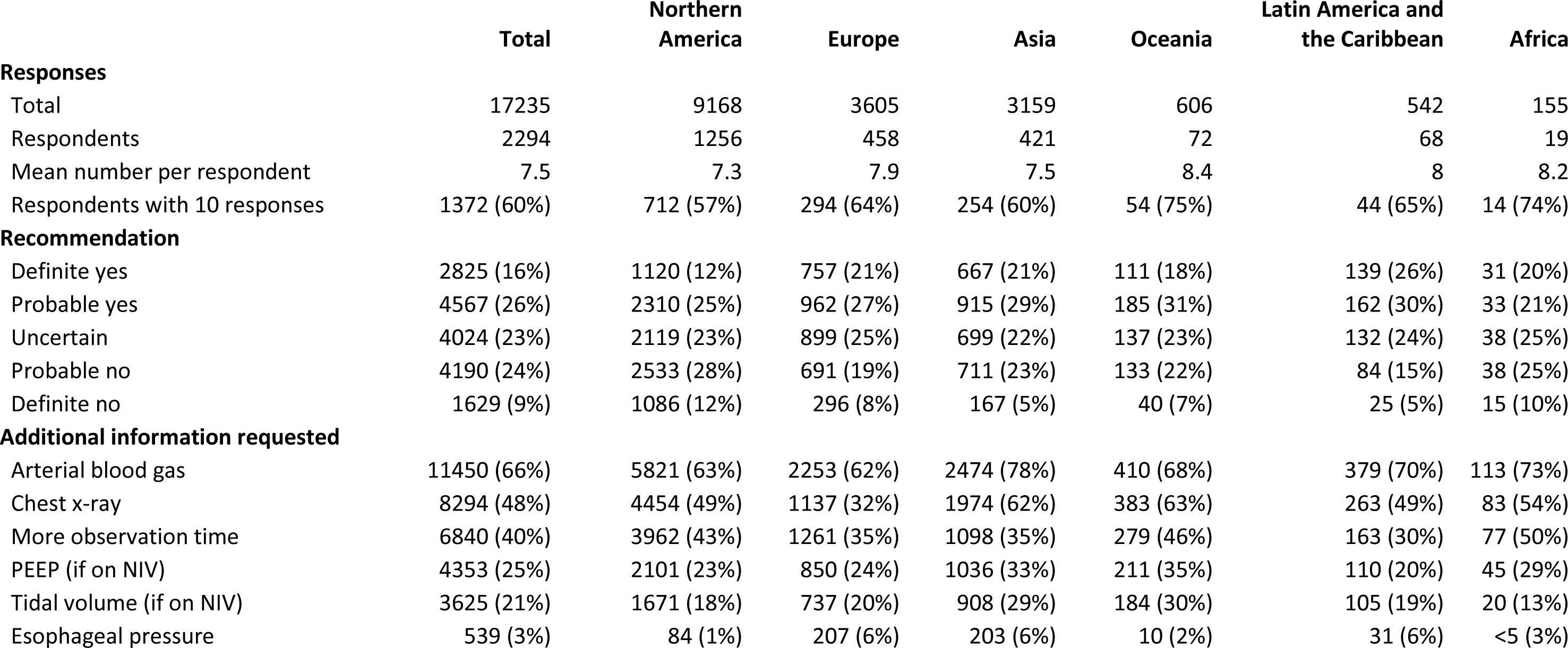

|  | Total | Northern<br>America | Europe | Asia | Oceania | Latin America and<br>the Caribbean | Africa |
| --- | --- | --- | --- | --- | --- | --- | --- |
| **Responses** |  |  |  |  |  |  |  |
| Total | 17235 | 9168 | 3605 | 3159 | 606 | 542 | 155 |
| Respondents | 2294 | 1256 | 458 | 421 | 72 | 68 | 19 |
| Mean number per respondent | 7.5 | 7.3 | 7.9 | 7.5 | 8.4 | 8 | 8.2 |
| Respondents with 10 responses | 1372 (60%) | 712 (57%) | 294 (64%) | 254 (60%) | 54 (75%) | 44 (65%) | 14 (74%) |
| **Recommendation** |  |  |  |  |  |  |  |
| Definite yes | 2825 (16%) | 1120 (12%) | 757 (21%) | 667 (21%) | 111 (18%) | 139 (26%) | 31 (20%) |
| Probable yes | 4567 (26%) | 2310 (25%) | 962 (27%) | 915 (29%) | 185 (31%) | 162 (30%) | 33 (21%) |
| Uncertain | 4024 (23%) | 2119 (23%) | 899 (25%) | 699 (22%) | 137 (23%) | 132 (24%) | 38 (25%) |
| Probable no | 4190 (24%) | 2533 (28%) | 691 (19%) | 711 (23%) | 133 (22%) | 84 (15%) | 38 (25%) |
| Definite no | 1629 (9%) | 1086 (12%) | 296 (8%) | 167 (5%) | 40 (7%) | 25 (5%) | 15 (10%) |
| **Additional information requested** |  |  |  |  |  |  |  |
| Arterial blood gas | 11450 (66%) | 5821 (63%) | 2253 (62%) | 2474 (78%) | 410 (68%) | 379 (70%) | 113 (73%) |
| Chest x-ray | 8294 (48%) | 4454 (49%) | 1137 (32%) | 1974 (62%) | 383 (63%) | 263 (49%) | 83 (54%) |
| More observation time | 6840 (40%) | 3962 (43%) | 1261 (35%) | 1098 (35%) | 279 (46%) | 163 (30%) | 77 (50%) |
| PEEP (if on NIV) | 4353 (25%) | 2101 (23%) | 850 (24%) | 1036 (33%) | 211 (35%) | 110 (20%) | 45 (29%) |
| Tidal volume (if on NIV) | 3625 (21%) | 1671 (18%) | 737 (20%) | 908 (29%) | 184 (30%) | 105 (19%) | 20 (13%) |
| Esophageal pressure | 539 (3%) | 84 (1%) | 207 (6%) | 203 (6%) | 10 (2%) | 31 (6%) | <5 (3%) |

Critical care medicine was the primary clinical area of practice for 1,825 (80%). Multiple primary areas were selected by 334 (15%), including anesthesia and emergency medicine. Most respondents were attending physicians (1437, 63%), and 507 (22%) respondents were nurses or respiratory therapists. Median duration of practice was 11 years (interquartile range [IQR] 5 to 20, eFigure 6).

### Responses

We gathered 17,235 vignette responses from the 2,294 respondents. 1,372 (60%) answered 10 scenarios (IQR 4 to 10, eFigure 7 & 8). Common responses included “Probable yes” (4,567, 26%), “Uncertain” (4,024, 23%), and “Probable no” (4,190, 24%). The remainder (22%) were either “Definite yes” or “Definite no.” Requests for additional information ranged from 11,450 (66%) for arterial blood gas to 539 (3%) for esophageal manometry.

### Primary analysis: Intubation recommendation

Several variables were associated with recommending intubation (Figure 1). Among clinician characteristics, a role of nurse or specialty of emergency medicine were associated with decreased odds of recommending intubation. Among patient characteristics, patient age, frailty, or diagnosis were not associated with recommending intubation. For oxygen saturation, inspired oxygen fraction, respiratory rate, breathing pattern, and level of consciousness, increasing derangement from normal was associated with increasing odds of recommending intubation (Figure 1). Only 2 of the 454 interaction term coefficients in the model had credible intervals not including 1, and in both cases this was likely artifactual due to limitations of an ordinal scale. (Supplement, section 8, eFigures 9 & 10).

**Figure 1:**
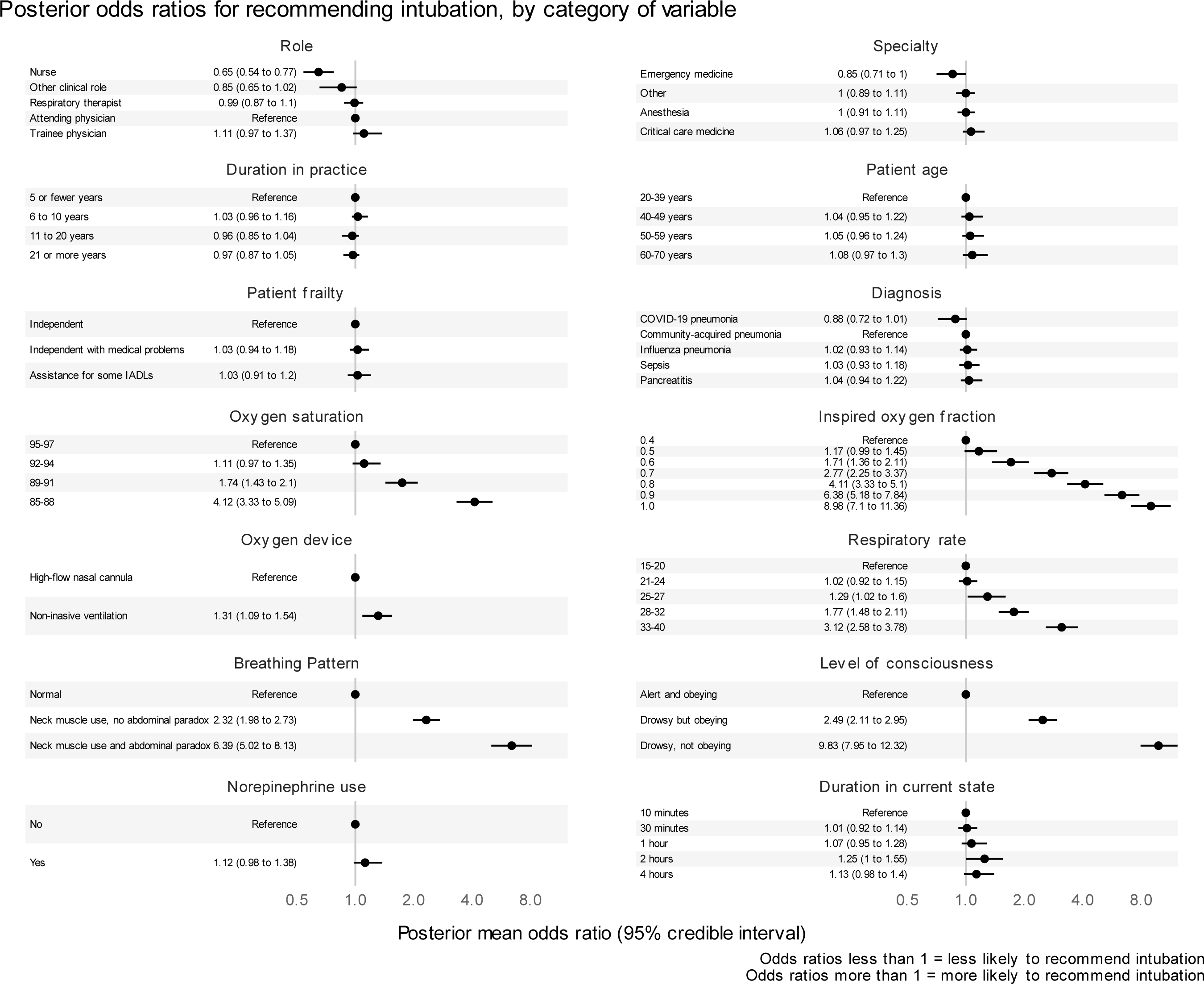
Posterior odds ratios for recommending intubation. Caption: This figure shows the posterior odds ratios across respondent, patient, and physiological variables for the strength of intubation recommendation (based on the ordinal response scale comprised of Definite no / Probable no / Uncertain / Probable Yes / Definite Yes). Odds ratios greater than 1 indicate variables associated with higher odds of recommending intubation, ie. a recommendation more towards the “Definite yes” end of the scale, whereas odds ratios less than 1 indicate variables associated with lower odds of recommending intubation, ie. a recommendation more towards the “Definite no” end of the scale.

### Variation by individual, country, and region

We quantified variability between individuals, countries, and regions using the median odds ratio.(41,42) The median odds ratio between individuals within the same country was 2.60 (CrI 2.48 to 2.73), indicating that randomly switching between individuals increased or decreased the odds of recommending intubation by an average factor of 2.6, after accounting for all other variables in the model. The median odds ratio between countries within the same region was 1.56 (CrI 1.35 to 1.85), and 1.17 (CrI 1.01 to 1.63) between regions.

The odds ratio for recommending intubation by country varied, but there was important uncertainty in these estimates for most countries (Figure 2). Only 6 of 74 countries had odds ratios with 95% credible intervals that did not include 1: Bangladesh (OR 0.39, CrI 0.20 to 0.74), Canada (OR 0.53, CrI 0.40 to 0.70), United States of America (OR 0.63, CrI 0.48 to 0.84), Japan (OR 1.37, CrI 1.02 to 1.86), Thailand (OR 1.48, CrI 1.03 to 2. 08), and Argentina (OR 2.23, CrI 1.31 to 3.94).

**Figure 2:**
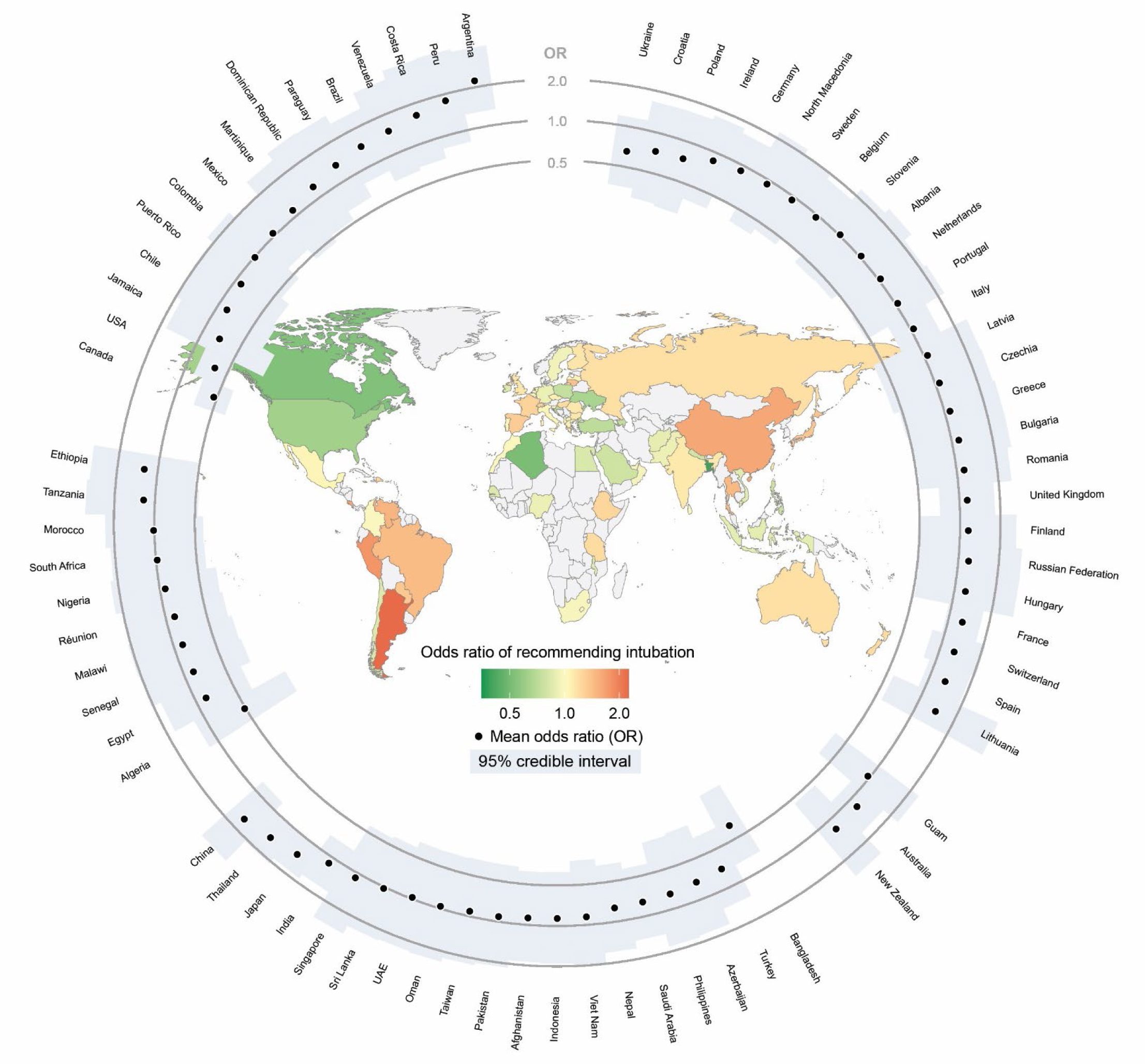
International variation in odds of recommending intubation. This figure shows the odds ratios for recommending intubation associated with respondent countries. The mean odds ratio is shown both as a black dot in the peripheral ring, and as the colour of the country within the map. The 95% credible interval or uncertainty or the estimate is shown by the size of the light blue bar in the peripheral ring. The odds ratios correspond to the combined random intercepts for region and country from the multilevel Bayesian proportional odds model. This means that countries with few respondents have wide uncertainty and their posterior estimates have been shrunk towards an odds ratio of 1.

### Secondary analysis: Additional information

Factors associated with requesting additional information varied by information type (Figure 3). No respondent or scenario variables were associated with requesting tidal volume or esophageal manometry. Nurse or respiratory therapist role was associated with increased odds of requesting arterial blood gas and chest x-ray, and respiratory therapist role was also associated with requesting PEEP information. Patient state of drowsy and not obeying was associated with increased odds of requesting an arterial blood gas, and decreased odds of requesting PEEP information or more observation time.

**Figure 3:**
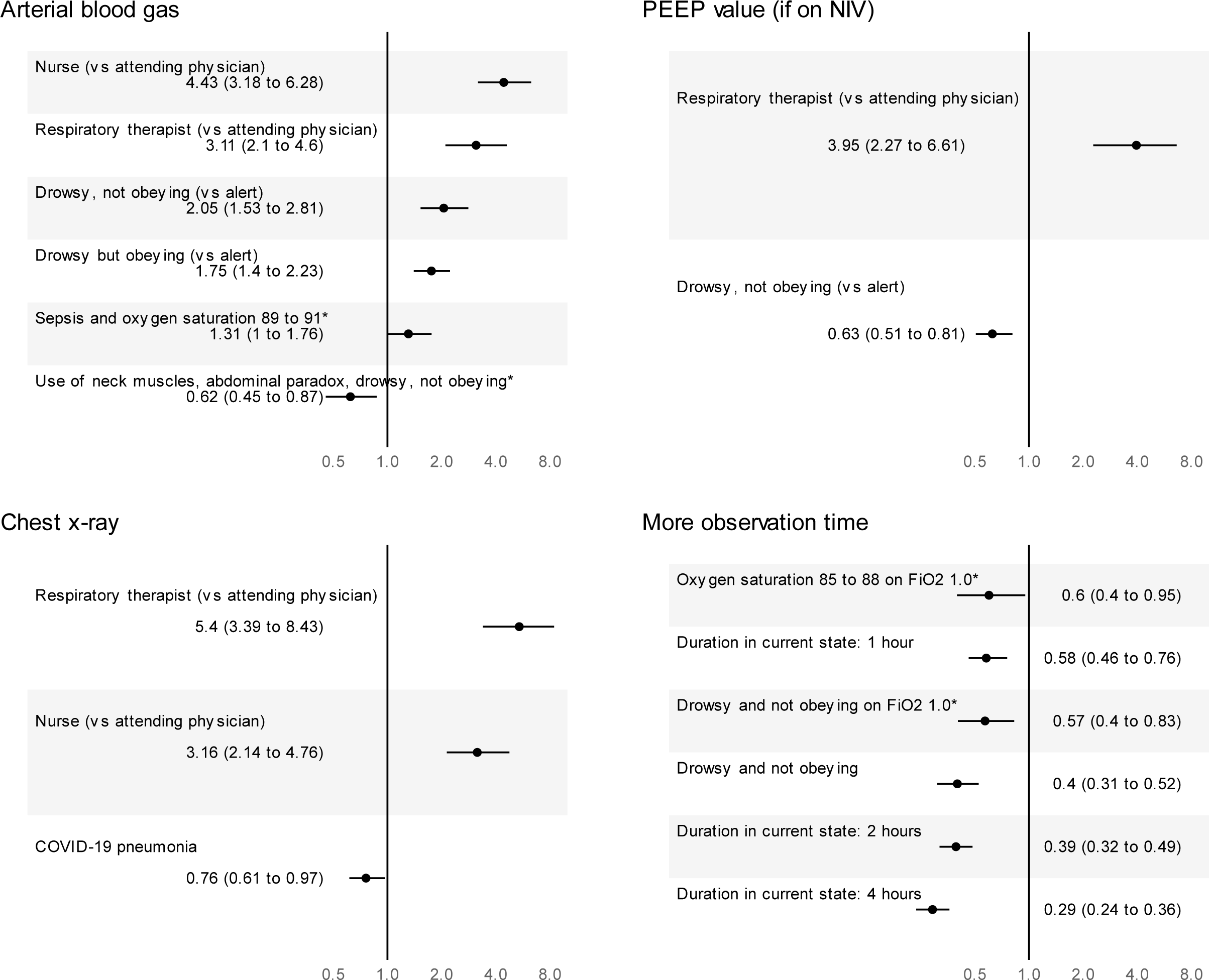
Posterior odds ratios for requesting additional information. This figure shows the posterior odds ratios associated with requesting each additional type of information. For clarity, we present only the posterior odds ratios with 95% credible intervals that do not include 1. For two types of additional information (tidal volume if on non-invasive ventilation, and esophageal manometry), there were no odds ratios that met this criterion. PEEP = positive end-expiratory pressure, NIV = non-invasive ventilation, COVID-19 = coronavirus 2019. *These odds ratios are interactions between the described variables.

### Secondary analysis: Comparison to observed data

There were 18,263 observations from 826 patients in the MIMIC-IV cohort. Intubation occurred within 3, 8, and 24 hours following 745 (4%), 1,375 (8%), and 2,635 (14%) observations. The average predicted probabilities of each response across all cohort observations were: “Definite no” – 25%, “Probable no” – 41%, “Uncertain” – 20%, “Probable yes” – 11%, “Definite yes” – 3%. The probability of “Definite yes” recommendations was similar to the observed intubation rate within 3 hours, and the probability of “Definite yes” or “Probable yes” recommendations was similar to the probability of observed intubation within 24 hours (eFigure 11). Predicted intubation recommendations of “Definite yes”, or “Definite yes and “Probable yes”, had similar discrimination with respect to intubation at 3 hours with an area under receiver operating curve of 0.71 (eFigures 12 & 13).

### Sensitivity analyses

The sensitivity analyses limited to respondents with 10 responses (eFigures 14 & 15) or divided by diagnosis had similar results to the primary analysis (eFigures 16-20). The proportional odds assumption was supported for all variables except nurse respondent role (eFigures 21-24).

## Discussion

In this survey of when to intubate patients with acute hypoxemic respiratory failure, we gathered 17,235 vignette-based intubation recommendations from 2,294 clinicians in 74 countries. Oxygenation, breathing pattern, and level of consciousness had a strong influence on intubation recommendations, while diagnosis, norepinephrine use, and duration of symptoms did not. We also found important variation in recommendations across clinicians and countries. The results persisted in multiple sensitivity analyses, and the survey responses had good calibration and moderate discrimination with respect to external data. Our findings suggest important elements of criteria to guide intubation, but also highlight uncertainty as to which criteria may be optimal in a given scenario.

We found that oxygenation, breathing pattern, and level of consciousness were strongly associated with intubation recommendations. These variables are highly relevant to the decision to intubate, as evidenced by their inclusion as intubation criteria in clinical trials(23,51–53) and by qualitative and survey data.(11–13) Our findings are notable for the magnitude of the odds ratios and the robust dose-response relationship between physiological derangement and probability of recommending intubation. For example, the probability of a respondent answering “Definite yes” or “Probable yes” increased from 33% to 53% after applying the odds ratio for use of neck muscles (2.32), and increased further to 76% after applying the odds ratio for use of neck muscles and abdominal paradox (6.39). The odds ratios relating to breathing pattern, inspired oxygen fraction, and level of consciousness were large enough that extreme values of any of these variables likely determine a clinician’s recommendation.

A respondent role of nurse was associated with modestly decreased odds of recommending intubation. Nurse respondents were also more likely to request arterial blood gases and chest x-rays. Potential explanations could include that nurses are not the final decision-maker, that nurses are less convinced of the benefits of invasive ventilation in a given scenario, or because higher rates of burnout in ICU nurses than ICU physicians(54) may contribute to a sense of therapeutic nihilism.

We identified several variables that were not associated with intubation recommendations, including diagnosis, norepinephrine use, duration in current state, and almost all of the 454 interaction terms in the model. The findings for diagnosis, where not even COVID-19 pneumonia was associated with decreased odds, suggest that intubation decisions are based on how a diagnosis affects oxygenation, breathing pattern, and level of consciousness, rather than on the diagnosis itself. Our findings are also consistent with other studies showing that norepinephrine use alone rarely determines intubation decision-making (16).

The lack of association between duration in current state or 99.5% of the interaction terms and intubation recommendations is somewhat surprising. Both clinical trajectory and the relationships between clinical variables have face validity as important components of intubation decision-making.(11) Although one explanation could be that “duration in current state” does not meaningfully capture clinical trajectory, our findings are consistent with cohort studies of patients with acute hypoxemic respiratory failure that suggest those who receive invasive ventilation are often those who fail to improve, and remain in the same (hypoxemic/ tachypneic) state over time, as opposed to those who worsen.(55,56) Regarding interactions, our findings suggest that clinicians gravitate towards parsimonious, heuristic approaches which do not rely on interactions.(57,58)

We found important variation in intubation recommendations between individual clinicians and countries. The median change in odds of recommending intubation between two individuals from the same country was 2.60, which was numerically similar to the change in odds between patients on inspired oxygen of 1.0 versus 0.7, patients with and without neck muscle use, or patients who are drowsy but obeying versus alert. These findings may partially explain prior work showing surprisingly low rates of intubation in ICU patients with AHRF who attain relatively severe degrees of physiologic derangement.(15)

While variation between countries was less marked than variation between individuals, odds ratios by country ranged from 0.39 to 2.23. A case-based survey of 1,136 intensivists focused on COVID-19 pneumonia management also found large variation in the proportion of respondents recommending intubation, ranging from 0% in Australia-New Zealand to 23% in Asia.(13) Our finding that clinicians in Canada and the USA were so much less likely to recommend intubation was striking, because health administrative data shows higher age-adjusted rates of invasive ventilation in both countries relative to the United Kingdom.(59) Potential explanations for country-level variation could relate to interpretation of the survey question, vignettes, and responses; access to resources including monitoring, non-invasive oxygen supports, and ventilators; or cultural, religious, and social practices and expectations regarding life support.

The most important limitation of our work is that a vignette-based survey is not guaranteed to reflect decisions made in clinical practice. Our secondary analysis using MIMIC-IV addresses this limitation, but provides only modest assurance that the survey responses match true clinical behaviour.

Selection bias due to non-response is another important limitation. We were unable to measure the response rate, which is one conventional way to assess vulnerability to selection bias. However, response rates in other surveys concerning intubation do not rule out the possibility of non-response bias(60), with rates ranging from 11% (12) to 50%(61). Surveys in other areas of critical care medicine have had response rates as low as 0.8%(62). Further, to calculate response rates, these surveys generally limited their distribution to medical societies with known membership. Our survey included non-physician clinicians involved in the decision to intubate, and physicians from countries and networks where the denominator is unknown. On balance, we felt that the benefits of a broader, larger sample more reflective of the global and team-based nature of intensive care medicine outweighed the potential harms of an unknown response rate.

Further limitations relate to survey design and modeling. The number of possible vignettes was much higher than the number of possible respondents, reflecting the multiplicative complexity of nuanced decision-making in medicine. This limited our ability to appreciate higher-order interactions. However, we obtained sufficient responses to evaluate bivariate interactions between any two variables. The proportional odds assumption implied by our modeling approach does not hold for all coefficients, based on our sensitivity analysis. However, proportional odds models can provide accurate treatment effect estimates even when there are deviations from the proportional odds assumption.(63–66) It was possible for respondents to complete the survey more than once, and so our primary analysis may be underestimating the individual-level variation. Reassuringly, our sensitivity analysis focused on those who fully completed the survey supported the primary analysis results.

## Conclusion

In this survey of 2,294 clinicians from 74 countries, intubation recommendations for patients with AHRF were associated with oxygenation, breathing pattern, and level of consciousness, and varied across individuals and countries. These data can help justify and inform randomized trials of physiological thresholds to guide intubation decisions that may improve patient-important outcomes.

## Data sharing statement

The data used in this study was deidentified and is available through reasonable request to the author. We plan to share the deidentified data through the Health Data Nexus repository (https://healthdatanexus.ai/) organized by the Temerty Center for Artificial Intelligence Research and Education in Medicine at the University of Toronto, in Toronto, Canada.

### Funding and role of funders in study

Survey dissemination costs were covered by an Interdepartmental Division of Critical Care Medicine Trainee Award to Dr Yarnell. Dr Fowler is the H. Barrie Fairley Professor of Critical Care at the University Health Network, Interdepartmental Division of Critical Care Medicine, University of Toronto. Funders had no role in the design and conduct of the study; collection, management, analysis, and interpretation of the data; preparation, review, or approval of the manuscript; nor in the decision to submit the manuscript for publication. The opinions, results and conclusions reported in this paper are those of the authors and are independent from the funding sources. No endorsement by any of the funding agencies is intended or should be inferred.

## Supporting information

Electronic supplement

## Data Availability

Deidentified data will be made available through Health Data Nexus

https://healthdatanexus.ai/

## Acknowledgements

We thank Dr Oleksa Rewa and Dr John Basmaji for comments on a draft of the manuscript. We thank the following for endorsement and/or dissemination: Australia New Zealand Intensive Care Society, Canadian Association of Critical Care Nurses, Canadian Association of Emergency Physicians, Canadian Critical Care Society, Canadian Society of Respiratory Therapists, the European Society of Intensive Care Medicine, Indian Registry of Intensive care and the Chennai branch of the Indian Society of Critical Care Medicine, Intensive Care Society (United Kingdom), Japanese Society of Respiratory Care, Pleural Pressure Working Group, Platform of Randomized Adaptive Clinical Trials in Critical Illness investigator group, Pragmatic Critical Care Research Group, REVA Network, Society for Critical Care Medicine, Societa’ Italiano di Anestesia, Analgesia, Rianimazione e Terapia Intensiva (SIAARTI), la Société de Réanimation de Langue Française (SRLF), and the Thai Society of Critical Care Medicine (TSCCM).

