## Supplementary material for "An international factorial vignette-based survey of intubation decisions in acute hypoxemic respiratory failure": Electronic supplement

Yarnell, Christopher J et al

2024-04-07

### 1 Contents

2 Supplement overview ... 3
3 EQUATOR CROSS Checklist ... 4
4 Survey design ... 7
4.1 eFigure 1: Survey preamble ... 9
4.2 eFigure 2: Survey demographics questionnaire ... 10
4.3 eFigure 3: Survey vignette question ... 11
4.4 eFigure 4: Survey additional information question ... 12
5 Vignette characteristics ... 13
5.1 eTable 1: Vignette characteristics ... 13
6 Survey dissemination ... 14
6.1 eFigure 5: Survey respondents by date and country ... 14
7 Additional figures ... 16
7.1 eFigure 6: Histogram of respondent duration in practice ... 16
7.2 eFigure 7: Number of responses per respondent ... 17
7.3 eFigure 8: Violin plot of time per scenario response by role ... 18
8 Primary analysis: additional details and results ... 19
8.1 Data preparation ... 19
8.2 Prior distributions ... 19
8.3 Model fit and diagnostics ... 19
8.4 Results: Interaction coefficients ... 20
8.5 eFigure 9: Probability of interaction odds ratios being greater than the region of practical equivalence (ROPE) 21
8.6 eFigure 10: Probability of interaction odds ratios being less than the region of practical equivalence (ROPE) .. 22
9 Comparison to observed data: MIMIC-IV cohort ... 24
9.1 Cohort construction ... 24
9.2 Comparing survey responses to observed data ... 25
9.3 eFigure 11: Calibration between quantiles of predicted probability of intubation recommendation and observed intubation rates ... 26
9.4 eFigure 12: Discrimination of probability of intubation recommendation of “Definite yes” and observed intubation ... 27
9.5 eFigure 13: Discrimination of probability of intubation recommendation of “Definite yes” or “Probable yes” and observed intubation ... 28
10 Sensitivity analysis of respondents with 10 responses ... 29
10.1 Median odds ratios ... 29
10.2 eFigure 14: Sensitivity analysis, forest plot of odds ratios for main effects ... 30
10.3 eFigure 15: Sensitivity analysis, forest plot of combined region and country-level odds ratios ... 31
11 Secondary analyses by patient diagnosis ... 32

### 2 Supplement overview

This supplement contains further information on the survey design, dissemination, uptake, and analysis. There are additional results for primary, secondary, and sensitivity analyses, mostly in the form of figures.

### 3 EQUATOR CROSS Checklist

| Section/topic | Item (1) | Item description | Reported on page # |
| --- | --- | --- | --- |
| **Title and abstract** | | | |
| Title and abstract | 1a | State the word “survey” along with a commonly used term in title or abstract to introduce the study’s design. | 1 |
| | 1b | Provide an informative summary in the abstract, covering background, objectives, methods, findings/results, interpretation/discussion, and conclusions. | Abstract |
| **Introduction** | | | |
| Background | 2 | Provide a background about the rationale of study, what has been previously done, and why this survey is needed. | Intro |
| Purpose/aim | 3 | Identify specific purposes, aims, goals, or objectives of the study. | Intro |
| **Methods** | | | |
| Study design | 4 | Specify the study design in the methods section with a commonly used term (e.g., cross-sectional or longitudinal). | Methods |
| Data collection methods | 5a | Describe the questionnaire (e.g., number of sections, number of questions, number and names of instruments used). | Methods |
| | 5b | Describe all questionnaire instruments that were used in the survey to measure particular concepts. Report target population, reported validity and reliability information, scoring/classification procedure, and reference links (if any). | Methods |
| | 5c | Provide information on pretesting of the questionnaire, if performed (in the article or in an online supplement). Report the method of pretesting, number of times questionnaire was pre-tested, number and demographics of participants used for pretesting, and the level of similarity of demographics between pre-testing participants and sample population. | Methods |
| | 5d | Questionnaire if possible, should be fully provided (in the article, or as appendices or as an online supplement). | Supplement |
| Sample characteristics | 6a | Describe the study population (i.e., background, locations, eligibility criteria for participant inclusion in survey, exclusion criteria). | Methods |
| | 6b | Describe the sampling techniques used (e.g., single stage or multistage sampling, simple random sampling, stratified sampling, cluster sampling, convenience sampling). Specify the locations of sample participants whenever clustered sampling was applied. | Methods |
| | 6c | Provide information on sample size, along with details of sample size calculation. | --- |
| | 6d | Describe how representative the sample is of the study population (or target population if possible), particularly for population-based surveys. | --- |
| Survey administration | 7a | Provide information on modes of questionnaire administration, including the type and number of contacts, the location where the survey was conducted (e.g., outpatient room or by use of online tools, such as SurveyMonkey). | Methods and supplement |
| | 7b | Provide information of survey’s time frame, such as periods of recruitment, exposure, and follow-up days. | Methods |
| | 7c | Provide information on the entry process:<br>–>For non-web-based surveys, provide approaches to minimize human error in data entry.<br>–>For web-based surveys, provide approaches to prevent “multiple participation” of participants. | Methods / sensitivity analysis / supplement |
| Study preparation | 8 | Describe any preparation process before conducting the survey (e.g., interviewers’ training process, advertising the survey). | --- |

| | | | |
|---|---|---|---|
| Ethical considerations | 9a | Provide information on ethical approval for the survey if obtained, including informed consent, institutional review board [IRB] approval, Helsinki declaration, and good clinical practice [GCP] declaration (as appropriate). | Methods |
| | 9b | Provide information about survey anonymity and confidentiality and describe what mechanisms were used to protect unauthorized access. | Methods |
| Statistical analysis | 10a | Describe statistical methods and analytical approach. Report the statistical software that was used for data analysis. | Methods |
| | 10b | Report any modification of variables used in the analysis, along with reference (if available). | Methods |
| | 10c | Report details about how missing data was handled. Include rate of missing items, missing data mechanism (i.e., missing completely at random [MCAR], missing at random [MAR] or missing not at random [MNAR]) and methods used to deal with missing data (e.g., multiple imputation). | Methods |
| | 10d | State how non-response error was addressed. | Methods |
| | 10e | For longitudinal surveys, state how loss to follow-up was addressed. | --- |
| | 10f | Indicate whether any methods such as weighting of items or propensity scores have been used to adjust for non-representativeness of the sample. | --- |
| | 10g | Describe any sensitivity analysis conducted. | Methods |
| **Results** | | | |
| Respondent characteristics | 11a | Report numbers of individuals at each stage of the study. Consider using a flow diagram, if possible. | Results |
| | 11b | Provide reasons for non-participation at each stage, if possible. | --- |
| | 11c | Report response rate, present the definition of response rate or the formula used to calculate response rate. | --- |
| | 11d | Provide information to define how unique visitors are determined. Report number of unique visitors along with relevant proportions (e.g., view proportion, participation proportion, completion proportion). | --- |
| Descriptive results | 12 | Provide characteristics of study participants, as well as information on potential confounders and assessed outcomes. | Results |
| Main findings | 13a | Give unadjusted estimates and, if applicable, confounder-adjusted estimates along with 95% confidence intervals and p-values. | Results |
| | 13b | For multivariable analysis, provide information on the model building process, model fit statistics, and model assumptions (as appropriate). | Results |
| | 13c | Provide details about any sensitivity analysis performed. If there are considerable amount of missing data, report sensitivity analyses comparing the results of complete cases with that of the imputed dataset (if possible). | Results |
| **Discussion** | | | |
| Limitations | 14 | Discuss the limitations of the study, considering sources of potential biases and imprecisions, such as non-representativeness of sample, study design, important uncontrolled confounders. | Discussion |
| Interpretations | 15 | Give a cautious overall interpretation of results, based on potential biases and imprecisions and suggest areas for future research. | Discussion |
| Generalizability | 16 | Discuss the external validity of the results. | Discussion |
| **Other sections** | | | |

| | | | |
|---|---|---|---|
| Role of funding source | 17 | State whether any funding organization has had any roles in the survey's design, implementation, and analysis. | Preamble |
| Conflict of interest | 18 | Declare any potential conflict of interest. | Preamble |
| Acknowledgements | 19 | Provide names of organizations/persons that are acknowledged along with their contribution to the research. | Preamble |

### 4 Survey design

We opted for a simple survey design (a single question) with randomized parameter values for the prespecified relevant variables. Several features of this design merit discussion.

The randomized element of the design had clear benefits. It allowed for samples over the entire multidimensional space of combinations of covariates, instead of sampling a small number of potential covariate combinations. Randomization of scenarios also prevented against measurement errors due to fatigue and question order.

The main drawback of the design was that we had to prespecify the variables included in the survey. It restricted our ability to discover new perspectives on recommending intubation which could not be captured by the setup of our scenarios. For this reason, we did include a field allowing for free text input on any other information that would be helpful for the respondent. We plan to analyze these data in a subsequent manuscript.

The randomization itself had some nuance, in order to ensure realistic vignettes.(2) With a simple randomization scheme using independent uniform distributions across the ranges of all variables, there were two many bizarre combinations of values. For example, seeing pancreatitis and community-acquired pneumonia at the same frequency was jarring, as was seeing respiratory rates of 20 and 40 with equal frequency. To smooth the experience of the respondent and make the scenarios more “life-like”, we changed the shapes of distributions and introduced covariance between variables.

Most of the changes in the shapes of distributions corresponded to increasing the probability of values in the center of the distribution. We set the probability of pancreatitis to be 1/9, and each of the other 4 diagnoses appeared with probability 2/9. The probability of “Independent with well-controlled comorbidities” was twice that of the other two frailty options. Oxygen saturations in the range of 90 to 92 had the highest probability, followed by 93-97 and 88-89, with 85-86 appearing least often. Respiratory rates in the 20-30 range were most common. Abdominal paradox was half as common as the other two breathing pattern options. “Alert and obeying” occurred twice as often as the other two level of consciousness options. Vasopressor use was more common if the diagnosis was sepsis and not allowed if the diagnosis was COVID. For patients less than 40, we only permitted the lowest frailty level, and we also did not allow the highest frailty level for patients from 40 to 55.

#### 4.1 eFigure 1: Survey preamble

### A survey of when to intubate in hypoxemic respiratory failure

Next

Thank you for your interest in our project! By completing this anonymous survey, you are consenting to our use of your responses for research. No identifying information, such as email addresses or IP addresses, is collected by the survey.

*This survey has been approved by the Research Ethics Board of the Scarborough Health Network in Toronto, Canada. Current version: v3_2023-08-15*

English | Español | Français | Português | Deutsch | Italiano | 官话 | 日本語 | ภาษาไทย | Bahasa Indonesia

**Instructions:**

1. Press next to record your responses and advance.

2. For each scenario, give your recommendation about intubation using the slider, then press 'Next.' Think carefully, because there is no going backwards once you press 'Next.'

3. You may answer up to 10 scenarios. The survey will disconnect and end if you are inactive for 10 minutes, but your responses up to that point will be recorded.

4. Assume that the patient would accept invasive ventilation if you felt it was indicated.

*Questions? Concerns? Want to collaborate? Contact the principal investigator Christopher Yarnell at*.*

#### 4.2 eFigure 2: Survey demographics questionnaire

Thank you for doing our survey.

**In your practice, are you involved in decisions about intubation for respiratory failure?**

- ○ Yes
- ○ No

**What is your clinical role?**

- ○ Attending physician
- ○ Trainee physician
- ○ Nurse
- ○ Respiratory therapist
- ○ Other clinical role
- ○ Non-clinical role

**What is your primary area of practice?**

- ○ Critical care medicine
- ○ Emergency medicine
- ○ Anesthesiology
- ○ Other

**How many years have you been in practice?**

5

**In which country do you practice?**

Canada ▾

☐ I'm not a robot

reCAPTCHA
Privacy - Terms

Next

#### 4.3 eFigure 3: Survey vignette question

Would you recommend intubation and invasive ventilation?

| Variable | Value |
|---|---|
| Diagnosis | Community-acquired pneumonia |
| Age (years) | 56 |
| Premorbid function | Independent with well-controlled medical problems |
| Peripheral oxygen saturation (%) | 96 |
| Inspired oxygen fraction | 0.8 |
| Oxygen device | High-flow nasal oxygen |
| Respiratory rate (breaths per minute) | 33 |
| Breathing pattern | Use of neck muscles, no abdominal paradox |
| Norepinephrine use | Yes |
| Level of consciousness | Drowsy but obeying |
| Duration in current state | 2 hours |

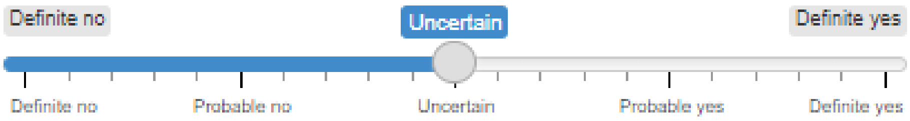

#### 4.4 eFigure 4: Survey additional information question

**Would any of the following additional information be useful to you?**

- Arterial blood gas
- PEEP (if on NIV)
- Tidal volume (if on NIV)
- Chest X-ray
- Esophageal pressure
- More observation time

**Any other information that would help you decide?**

Next

### 5 Vignette characteristics

#### 5.1 eTable 1: Vignette characteristics

| Variable | Possible values |
|---|---|
| Diagnosis | 1. Community-acquired pneumonia<br>2. COVID pneumonia<br>3. Influenza pneumonia<br>4. Pancreatitis<br>5. Sepsis |
| Age | 20 to 70 years |
| Frailty | 1. Independent and fit (CFS 1-2)<br>2. Independent with well-controlled medical problems (CFS 3)<br>3. Assistance for shopping and heavy housework (CFS 5) |
| Peripheral oxygen saturation | 85% to 97% |
| Inspired oxygen fraction | 0.4 to 1.0 in increments of 0.1 |
| Oxygen device | 1. High-flow nasal oxygen<br>2. Non-invasive ventilation |
| Respiratory rate | 15 to 40 breaths per minute |
| Work of breathing | 1. No use of neck muscles, no abdominal paradox<br>2. Use of neck muscles, no abdominal paradox<br>3. Use of neck muscles, abdominal paradox |
| Norepinephrine use | 1. No<br>2. Yes |
| Level of consciousness | 1. Alert and obeying<br>2. Drowsy but obeying<br>3. Drowsy and not obeying |
| Duration of abnormalities | 1. 10 minutes<br>2. 30 minutes<br>3. 1 hour<br>4. 2 hours<br>5. 4 hours |

Based on clinical sensibility testing, we removed patient ages greater than 70 and clinical frailty scales greater than 5, to reduce noise relating to differences in decision-making about invasive ventilation in older and more frail patients.(3,4) We also specified in the preamble that “the patient would accept invasive ventilation if the clinician felt it was indicated”, to reduce noise relating to uncertainty about patient preferences.(5)

Non-COVID-19 diagnoses were based on the distribution of diagnoses in a large, pre-COVID-19 randomized trial of non-intubated patients with acute hypoxemic respiratory failure.(6)

### 6 Survey dissemination

#### 6.1 eFigure 5: Survey respondents by date and country

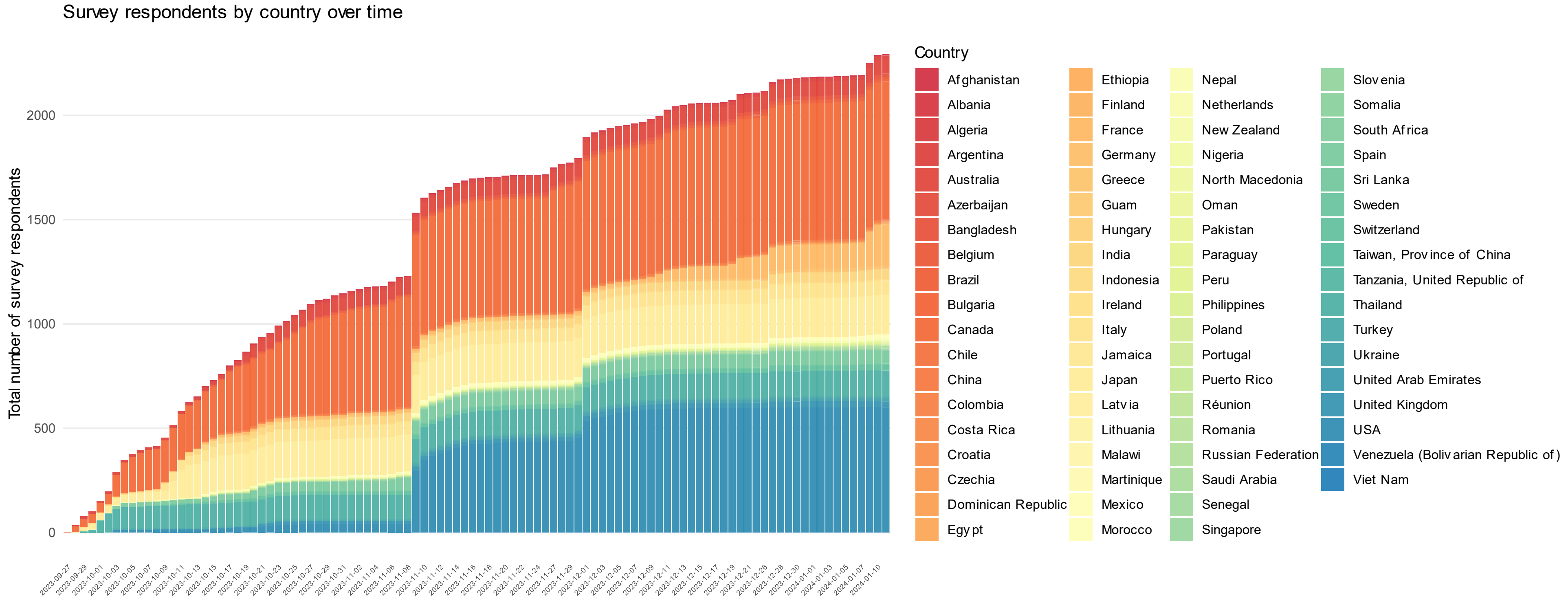

Caption: This figure shows recruitment over time by country.

Notable dates below.

- September 27, 2023: survey sent out to Canadian Critical Care Society (see Canadian responses, orange) and Thai intensive care networks (turquoise)
- October 11, 2023: survey sent out through Italian and Indian networks, and through Japanese Intensive Care Society (see Japanese responses, yellow)
- November 9, 2023: survey sent to Society for Critical Care Medicine (first round). Note large bump in USA respondents (blue)

- December 1, 2023: survey sent to Society for Critical Care Medicine (second round)
- Mid-December, 2023: survey publicized through REVA Network and la Société de Réanimation de Langue Française with corresponding increase in French respondents (yellow-orange)

### 7 Additional figures

#### 7.1 eFigure 6: Histogram of respondent duration in practice

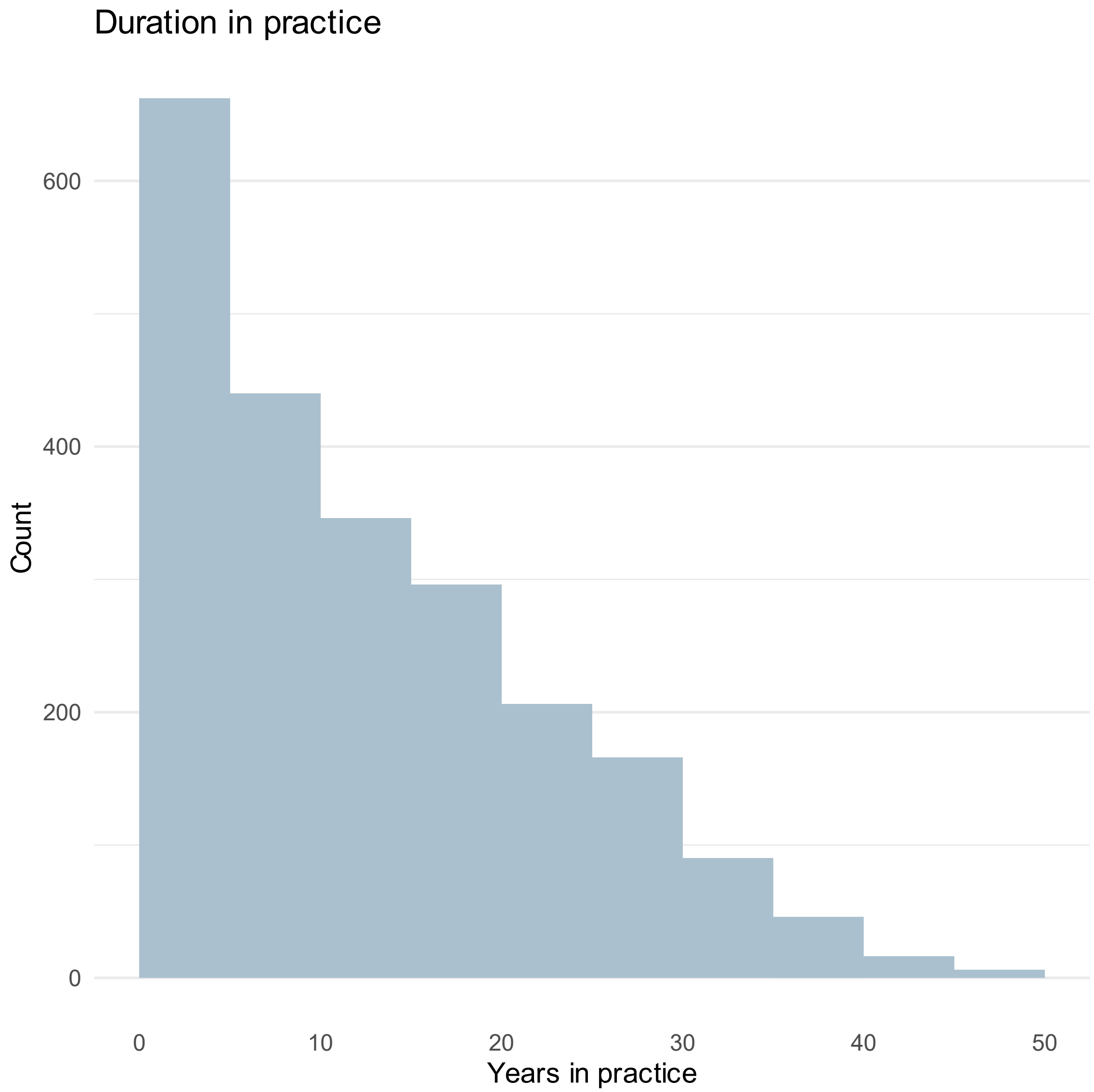

#### 7.2 eFigure 7: Number of responses per respondent

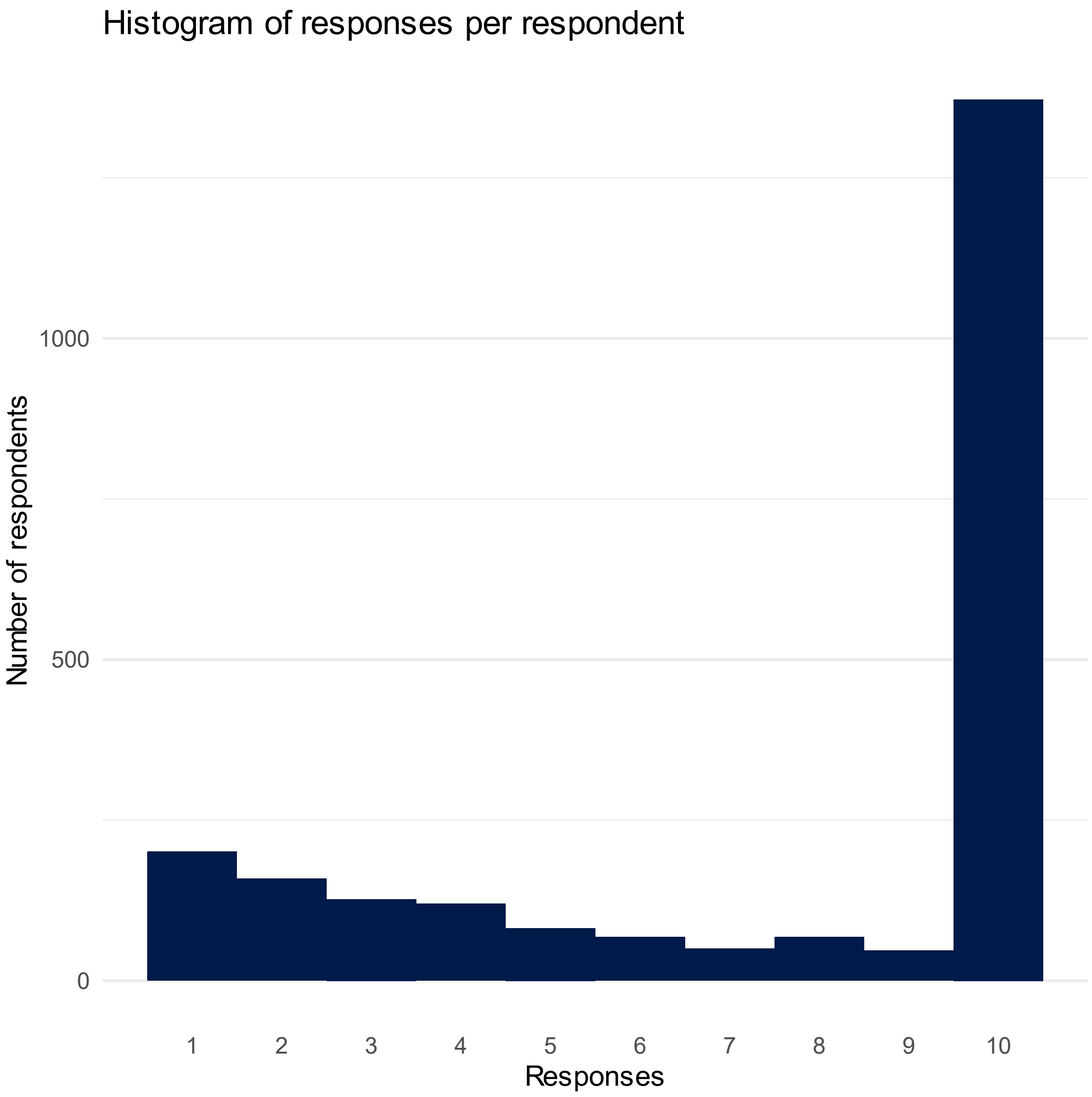

#### 7.3 eFigure 8: Violin plot of time per scenario response by role

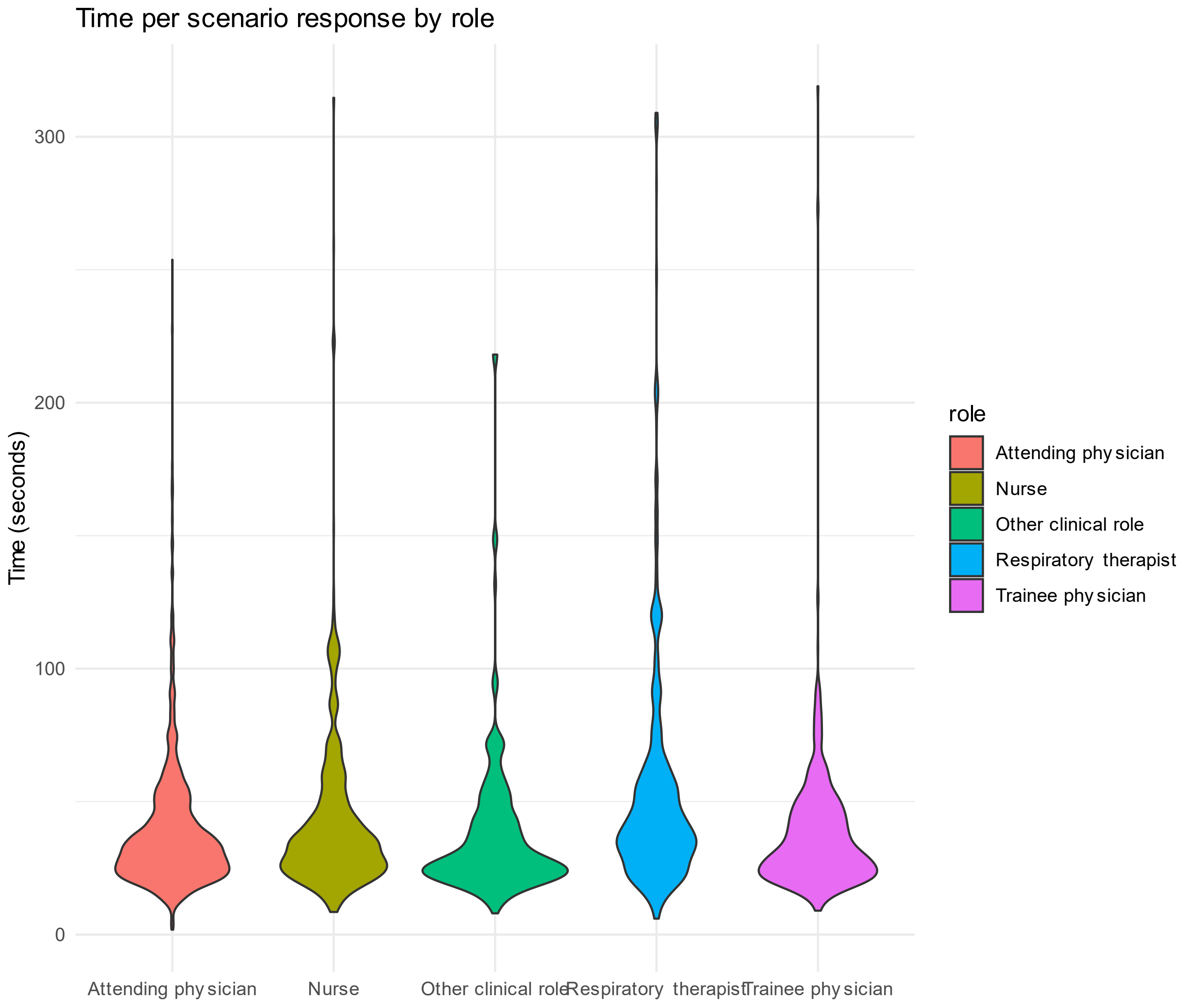

Median time per scenario was 33 seconds (IQR 24 to 46).

### 8 Primary analysis: additional details and results

#### 8.1 Data preparation

We categorized continuous variables in order to allow for non-linearity while also maintaining interpretable model results. We categorized variables as follows:

a. Respondent: specialty, role, duration in practice (0-5 years, 6-10 years, 11-20 years, more than 20 years)
b. Patient baseline information: age (20-39, 40-49, 50-59, 60-70), frailty, diagnosis
c. Patient clinical status: peripheral oxygen saturation (< 89, 89-91, 92-94, 95-97), inspired oxygen fraction (categorical), oxygen device, respiratory rate (<21,21-24, 25-27, 28-32, >32), breathing pattern, norepinephrine use, level of consciousness, duration in current state

#### 8.2 Prior distributions

Here we describe the prior distributions for the Bayesian proportional odds model used in the analyses. For all fixed effects (including interactions) we used a horseshoe prior distribution (3 degrees of freedom, 15% non-zero parameters). This prior belief holds that intubation decisions are parsimonious, that is, no more than 15% of the 501 parameters will have odds ratios for recommending intubation that are different from one.(7,8) See supplement for details regarding priors for random effects.

The random effects for clustering were modeled as normal distributions in the log-odds space with mean 0. The prior for the standard deviation of the random effects was a half-normal distribution with standard deviation 0.5.(9,10)

#### 8.3 Model fit and diagnostics

The primary model fit took approximately 6 hours on a 16GB laptop using 4 chains, each parallelized across 1 core. There were 2000 posterior samples with 3 divergent transitions. The rhat values were all less than 1.02, with 99% less than 1.01. Adapt_delta was set to 0.8 for the initial model run. We did re-run the model with adapt_delta set to 0.9 in order to assess if the divergent transitions indicated a biased posterior(11), and the results were very similar except that the model took 8 hours to run. There was only 1 divergent transition among the 2000 posterior samples. In the manuscript and supplement we present the results from the second model run, with adapt_delta set to 0.9.

#### 8.4 Results: Interaction coefficients

Of the 454 interaction terms in the model, all but 22 (5%) had posterior mean odds ratios within the region of practical equivalence (0.9 to 1.1). (eFigures 10 and 11) Of those 22, all but 2 had credible intervals overlapping 1. Those 2 were: saturation 85-88% and drowsy but not obeying (OR 0.76, CrI 0.61 to 0.96), and using neck muscles with abdominal paradox and drowsy but not obeying (OR 0.77, CrI 0.62 to 0.97). Both of these interaction terms are more likely to be a reflection of thresholding due to the limitations of an ordinal outcome than a true representation of clinical judgment. The odds ratios in question were: saturation 85-88% 4.12, drowsy not obeying 9.83, neck muscles and abdominal paradox 6.39. The interaction means that the odds ratio of intubation for a patient with saturation 85-88% and drowsy but not obeying was 30.8, not 40.5. For a baseline intubation recommendation probability of 20%, the final probability of recommending intubation would be 89% (with interaction) or 91% (without interaction). Similarly, the interaction means that the odds ratio of intubation for a patient that is drowsy but not obeying and is using neck muscles with abdominal breathing was 48.4, not 62.8. For a baseline intubation recommendation probability of 20%, the final probability of recommending intubation would be 92% (with interaction) or 94% (without interaction). These differences are not clinically significant, and instead they are most likely an artifact of the fact that a patient with any of those three variables (85-88%, drowsy and not obeying, using neck muscles with abdominal paradox) has a high probability of having "Definite yes" selected as the response, and there isn't enough room left in the ordinal scale to see a further increase when another of those variables is present.

#### 8.5 eFigure 9: Probability of interaction odds ratios being greater than the region of practical equivalence (ROPE)

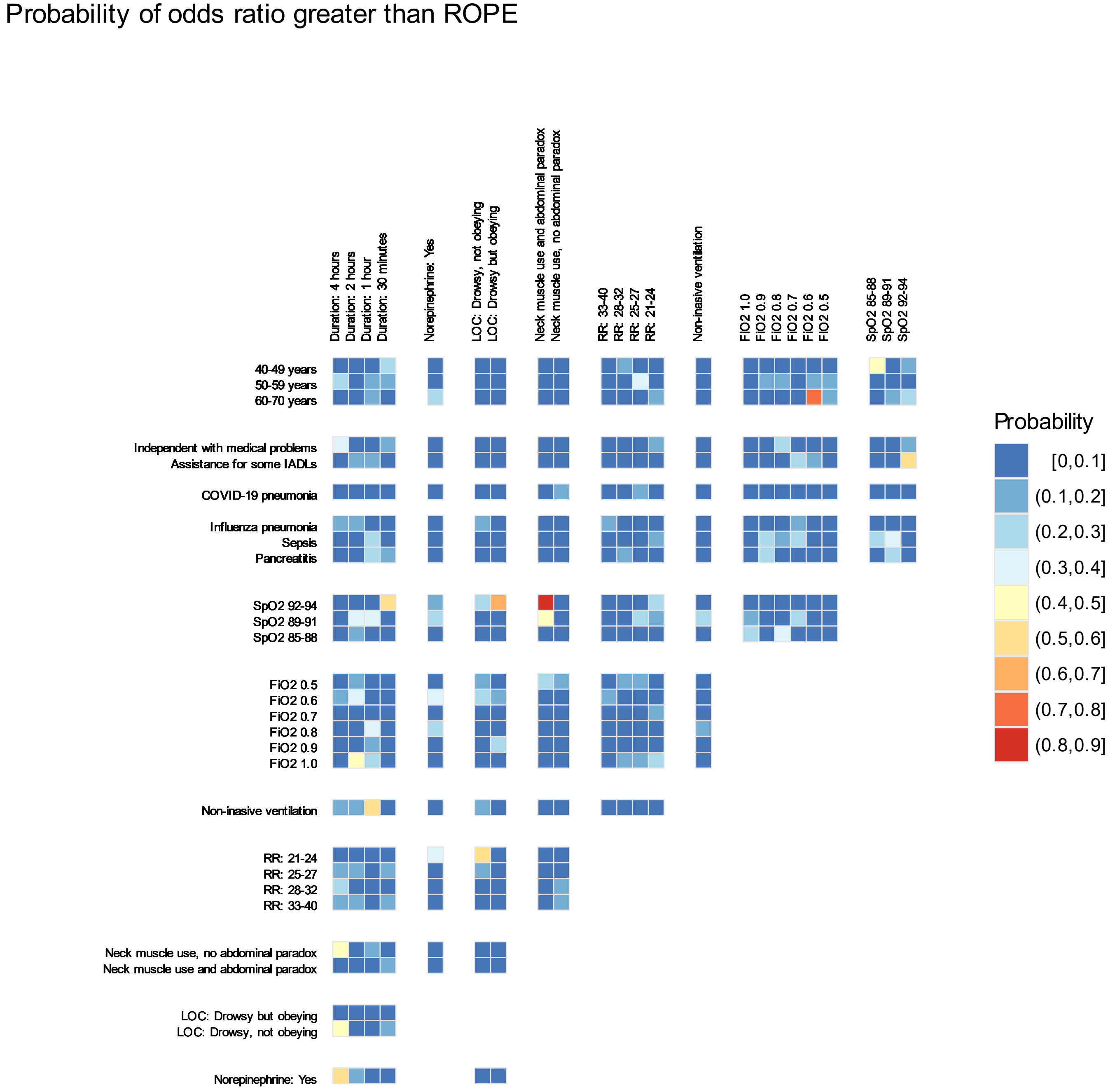

Caption: This figure shows a heat map of the probability that an interaction term was greater than the region of practical equivalence (that is, OR > 1.1). If the odds ratio is greater than the region of practical equivalence, that means that the combination of the two characteristics interacting were associated with a greater likelihood of recommending intubation than would have been expected by multiplying the odds ratios of each of the two characteristics. No interactions were greater than the region of practical equivalence with probability more than 0.9.

#### 8.6 eFigure 10: Probability of interaction odds ratios being less than the region of practical equivalence (ROPE)

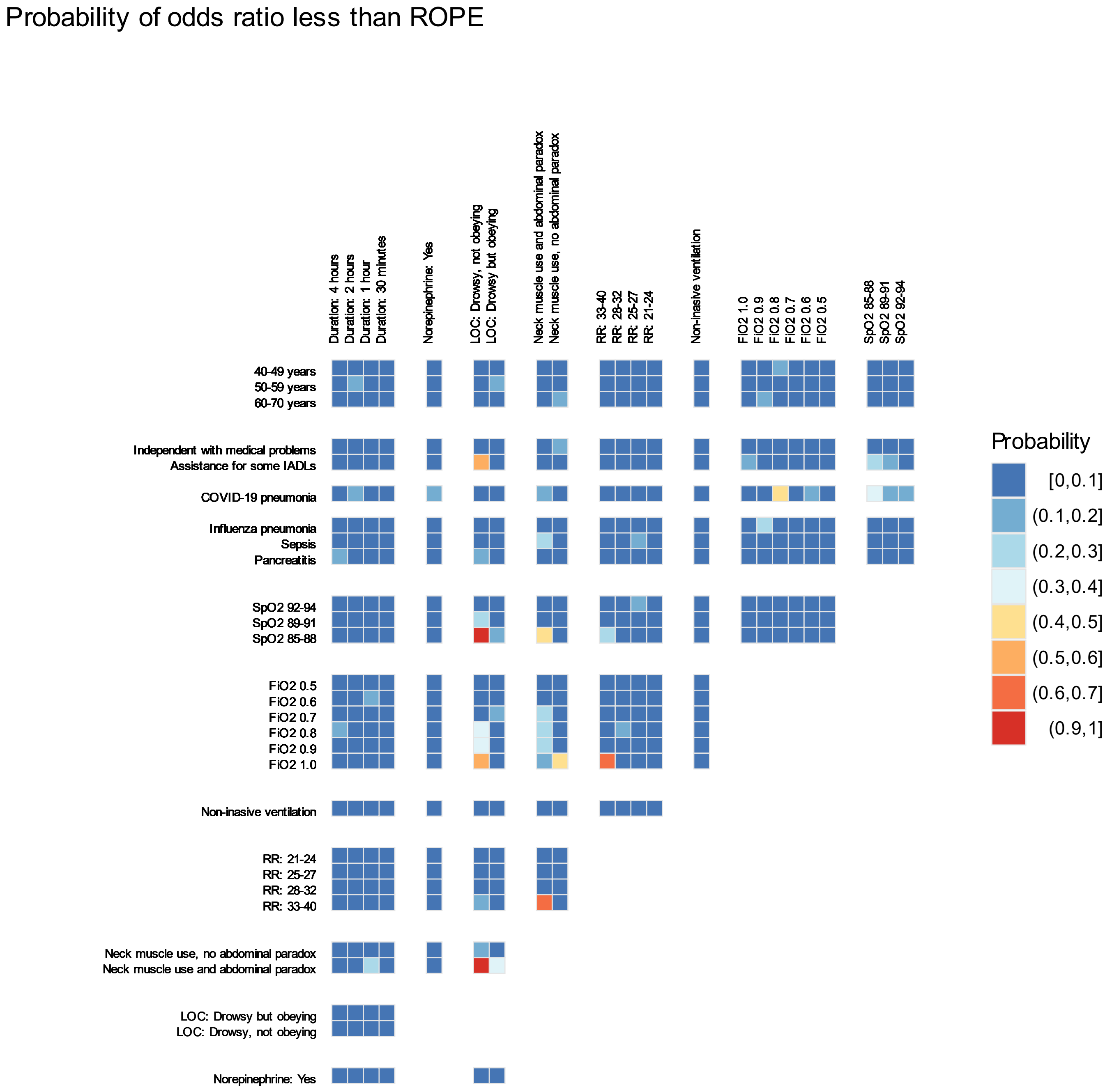

Caption: This figure shows a heat map of the probability that an interaction term was less than the region of practical equivalence (that is, OR < 0.9). If the odds ratio is less than the region of practical equivalence, that means that the combination of the two characteristics interacting were associated with a lower likelihood of recommending intubation than would have been expected by multiplying the odds ratios of each of the two characteristics. Both red tiles relate to a decreased level of consciousness (drowsy, not obeying) combined with either SpO2 85-88% or neck muscle use with abdominal paradox. As explained previously, the interaction slightly reduces the odds of recommending intubation in these scenarios, most likely because the probability of recommending intubation for each of the individual characteristics was already so high that the ordinal scale lacked resolution to show an increase that fully reflected the combination of the pairs of characteristics.

### 9 Comparison to observed data: MIMIC-IV cohort

#### 9.1 Cohort construction

The cohort was constructed in the same manner as described previously.(12) In brief, the cohort is built from the Medical Information Mart for Intensive Care version IV (MIMIC-IV).(13,14) We included patients who were admitted to the intensive care unit (ICU) at the Beth Israel Deaconess Medical Centre in Boston, USA. We included only patients who were receiving oxygen by non-rebreather mask, high-flow nasal cannula, or non-invasive ventilation with an inspired oxygen fraction of 0.4 or more within the first 24 hours of ICU admission.

From the cohort constructed for our prior investigations, we further filtered out all patients and patient observations which were not compatible with our model based on survey responses. As an example, in our survey, the maximum age was 70 years. Therefore we filtered out all patients who had age greater than 70 years. Thus we were left with observations from patients of age 20-70 years with saturations ranging from 85% to 97%, receiving oxygen by high-flow nasal cannula or non-invasive ventilation with an inspired oxygen fraction of 0.4 or more, with respiratory rate between 15 and 40 breaths per minute.

We also had to make some assumptions to render the MIMIC-IV data compatible with our survey model. We assumed that the responding clinician was an attending physician, specializing in critical care medicine, based in the USA, who had been practicing for 6-11 years. We assumed that all patients were “independent and fit” prior to admission, and that all patients had community-acquired pneumonia, given the limited difference between diagnoses (except for COVID, which was not present in the pre-pandemic database). We assumed that patients with an abnormal “work of breathing” variable had “Neck muscle use without abdominal paradox.” We converted the Glasgow Coma Scale observations as follows: GCS 15 = “Alert and obeying”, GCS 10-14 = “Drowsy but obeying”, GCS < 10 = “Drowsy, not obeying.” We rounded all inspired oxygen fractions to the nearest 0.1. We assumed that the duration of current abnormalities had been 1 hour, which is approximately the median period between observations in this dataset.

At the end we were left with 18,263 observations from 826 patients. For each of these recommendations, we used our model to predict how a survey respondent would have responded to a scenario with the same characteristics. The model predicted the probability that a response would take each of the available ordinal outcome values. For example, the model might have predicted as follows: Definite no – 10%, Probable no – 15%, Uncertain – 20%, Probable yes – 40%, Definite yes – 15%.

#### 9.2 Comparing survey responses to observed data

We compared predicted intubation recommendations of “Definite yes” and “Definite yes” or “Probable yes” to observed intubation rates within 3, 8, and 24 hours of each eligible observation. We reported the area under the receiver operating curve and calibration for each of the six pairs of predicted recommendation and intubation outcome.

#### 9.3 eFigure 11: Calibration between quantiles of predicted probability of intubation recommendation and observed intubation rates

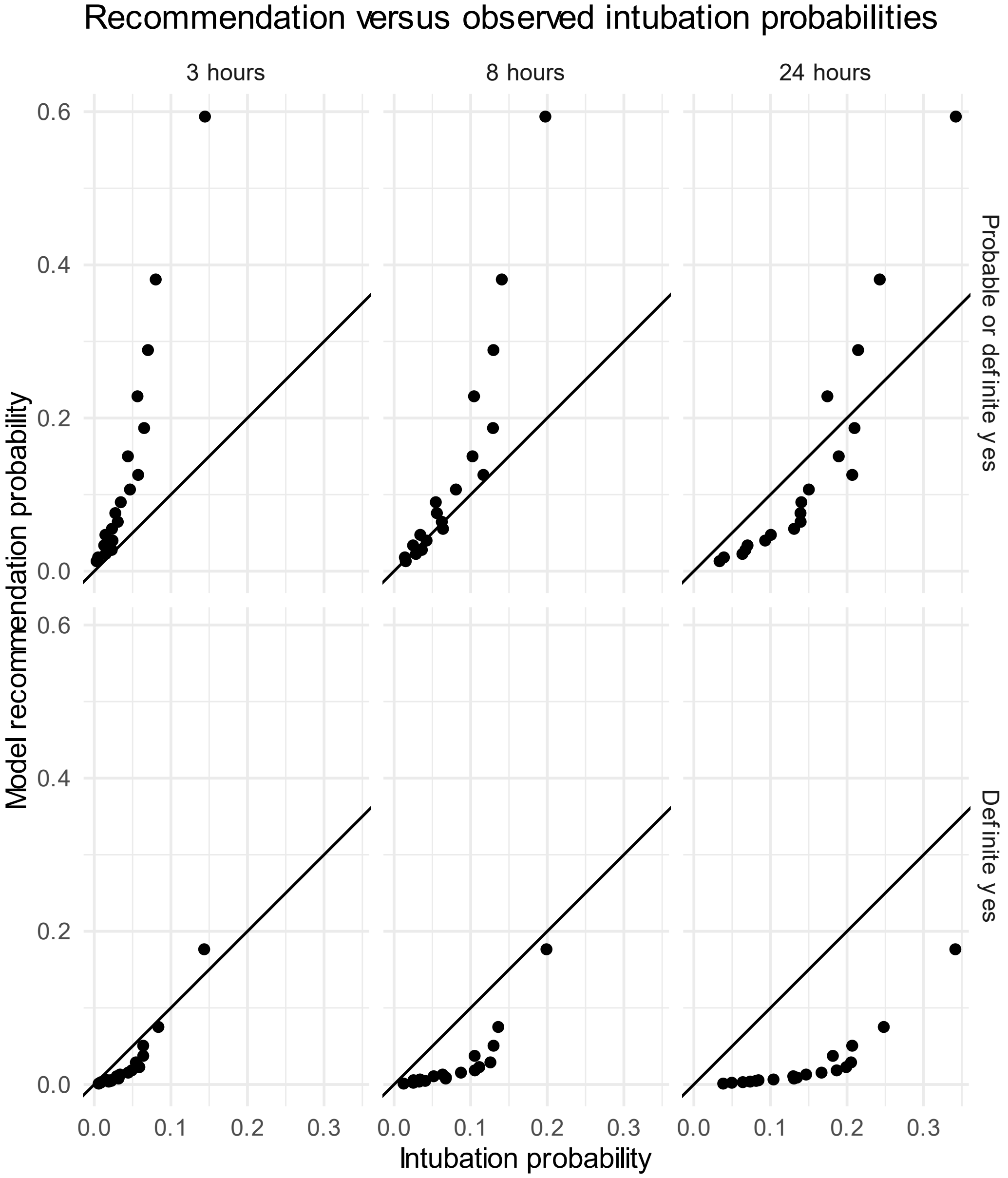

Figure compares 20 probability quantiles for observed intubation within a given time (x, columns) and probability of recommendation (y, rows). Observed cohort based on 18,263 observations from 826 patients from MIMIC-IV. Model recommendation probability based on Bayesian proportional odds model from survey.

#### 9.4 eFigure 12: Discrimination of probability of intubation recommendation of "Definite yes" and observed intubation

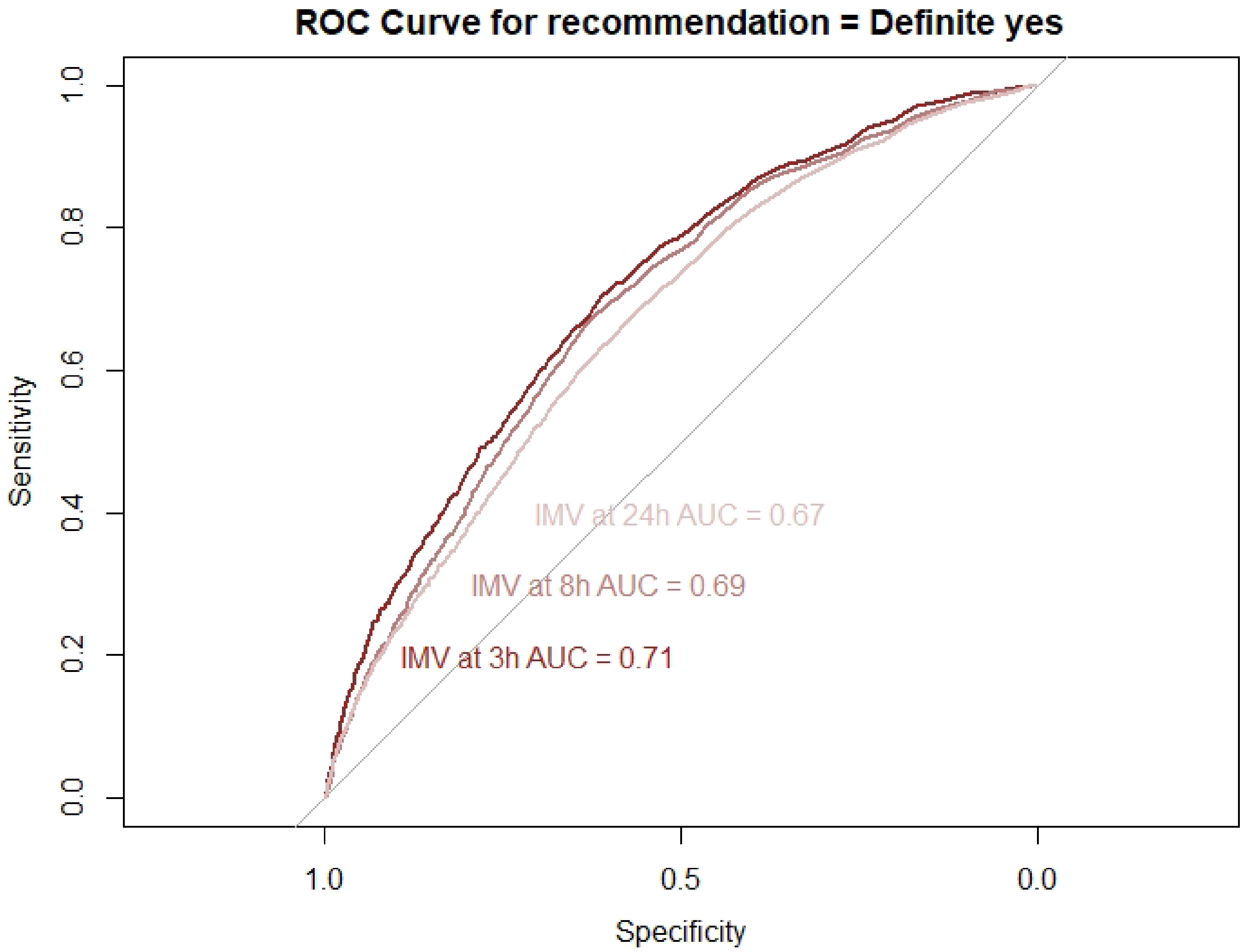

#### 9.5 eFigure 13: Discrimination of probability of intubation recommendation of “Definite yes” or “Probable yes” and observed intubation

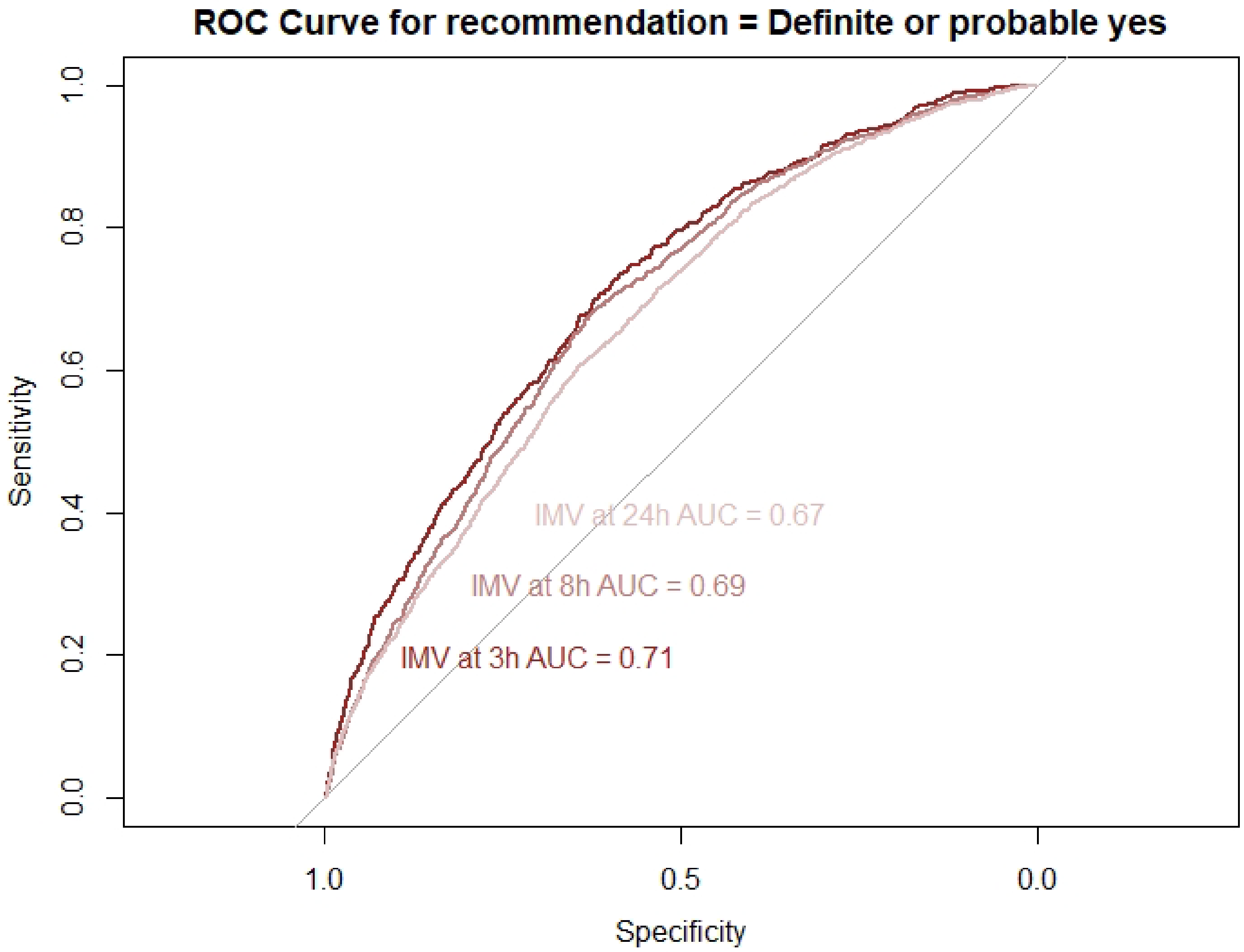

### 10 Sensitivity analysis of respondents with 10 responses

#### 10.1 Median odds ratios

The median odds ratio for switching between individuals within the same country was 2.60 (CrI 2.53 to 2.67), for switching between countries within the same region it was 1.88 (CrI 1.67 to 2.12), and for switching regions it was 1.46 (CrI 1.09 to 2.09).

#### 10.2eFigure 14: Sensitivity analysis, forest plot of odds ratios for main effects

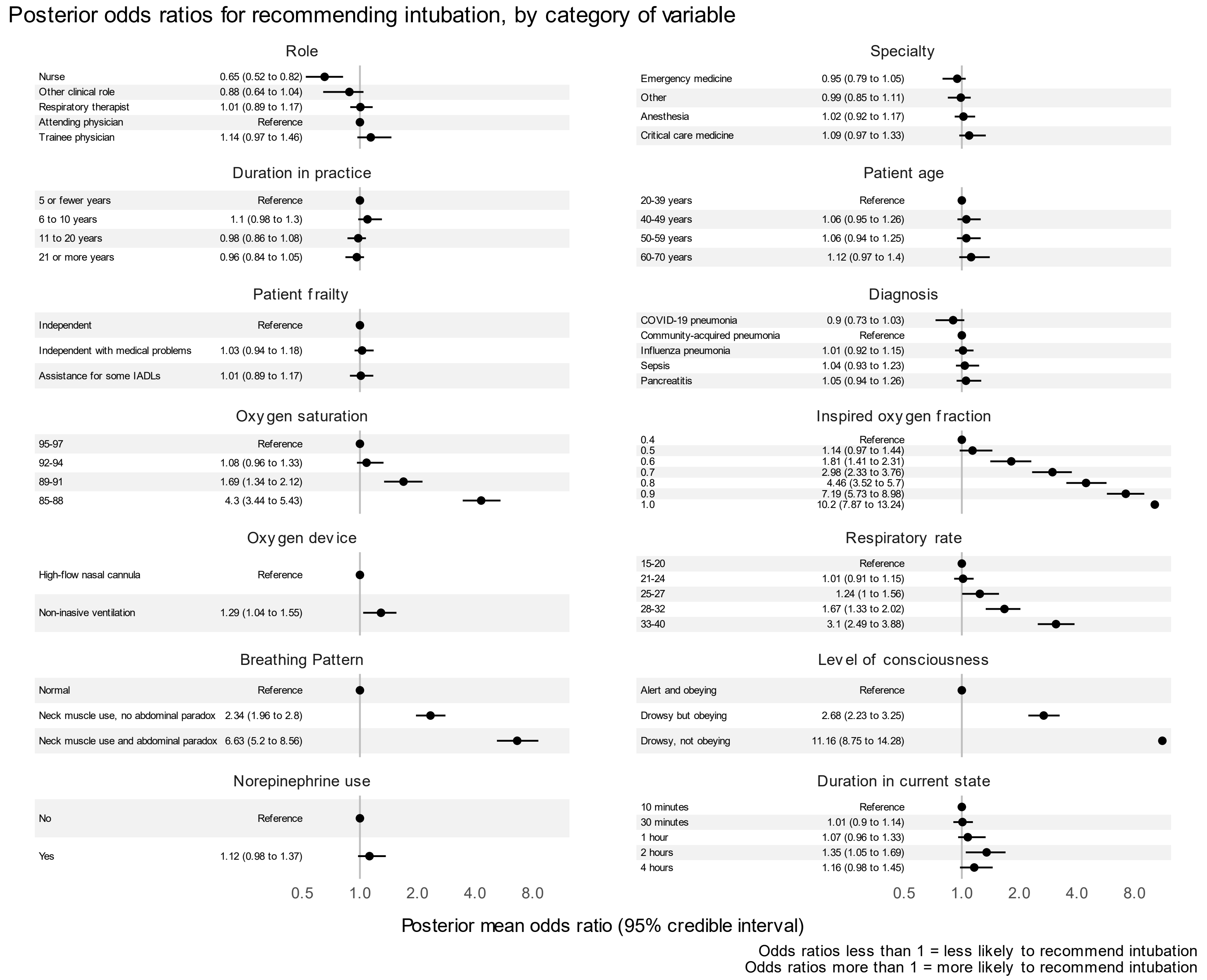

#### 10.3 eFigure 15: Sensitivity analysis, forest plot of combined region and country-level odds ratios

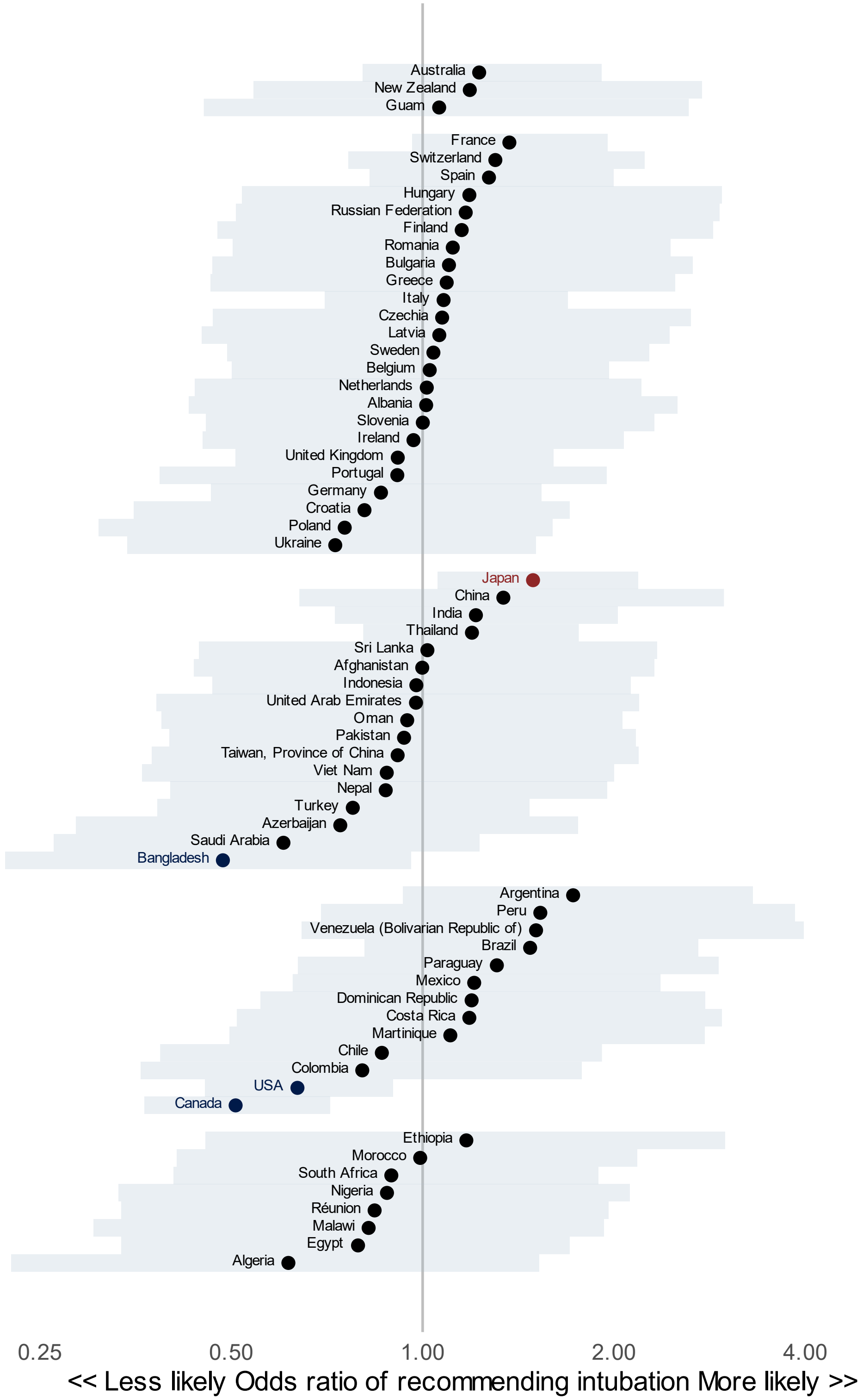

### 11 Secondary analyses by patient diagnosis

#### 11.1 eFigure 16: Posterior odds ratios, sepsis scenarios

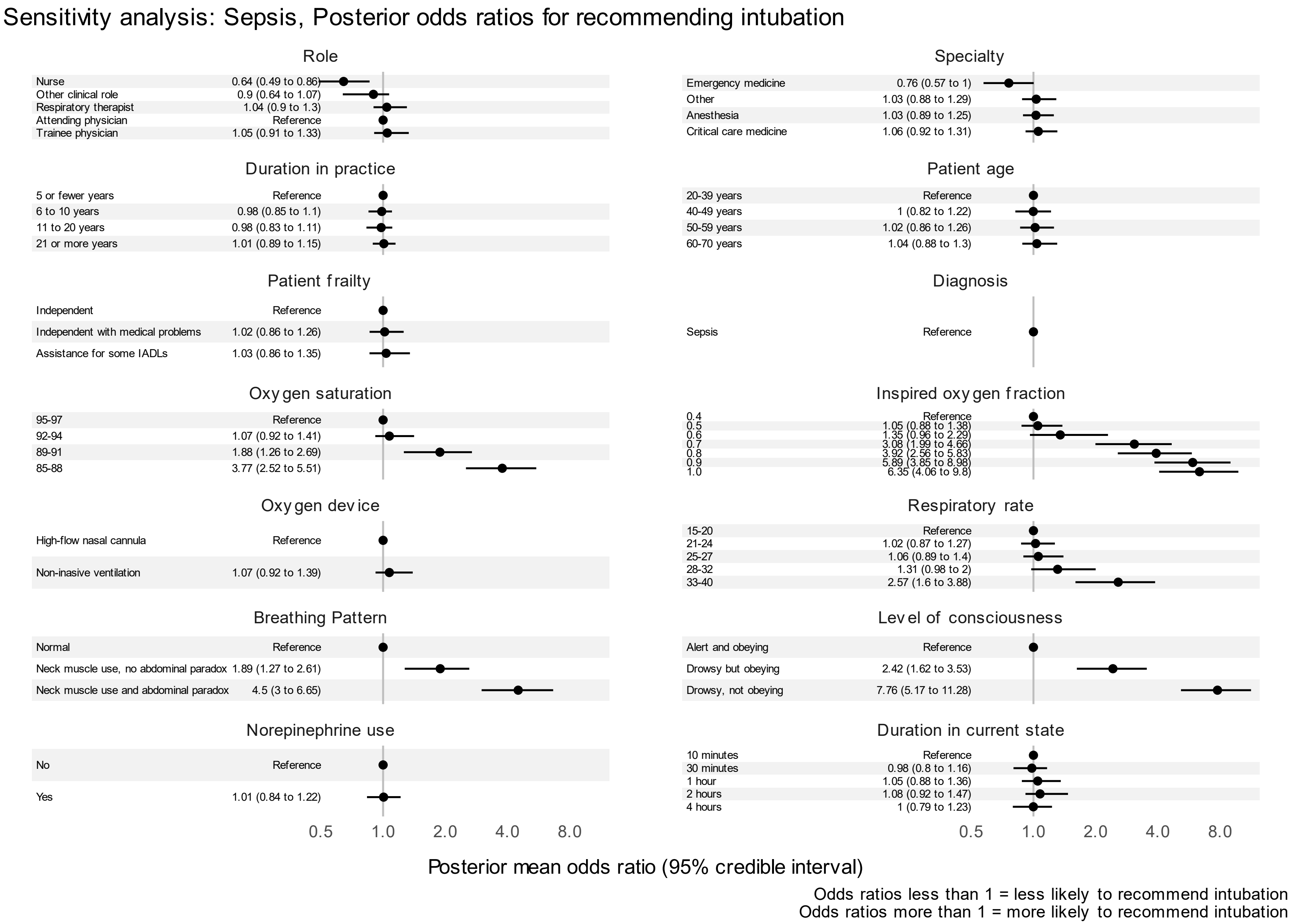

#### 11.2eFigure 17: Posterior odds ratios, community-acquired pneumonia scenarios

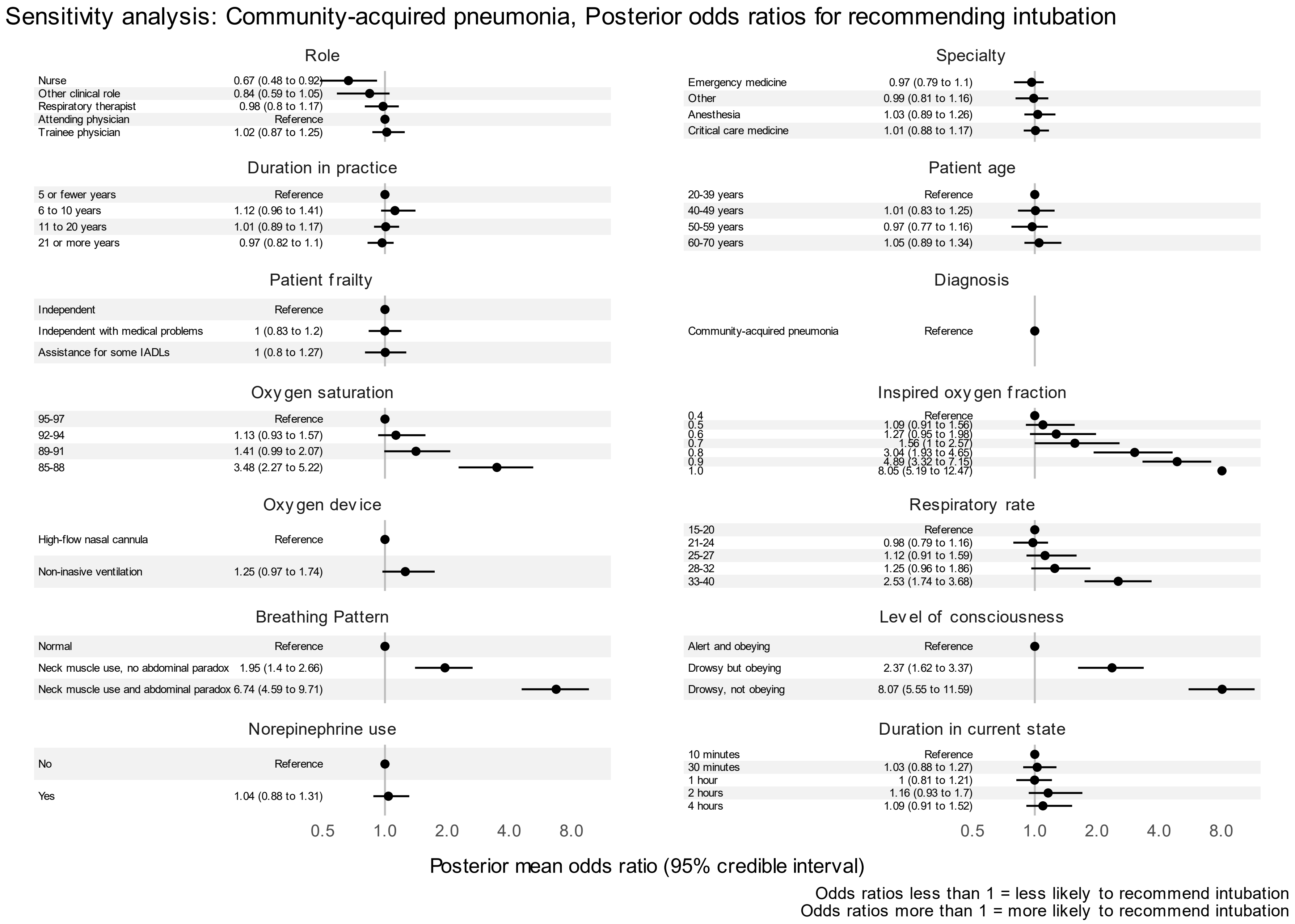

#### 11.3eFigure 18: Posterior odds ratios, influenza pneumonia scenarios

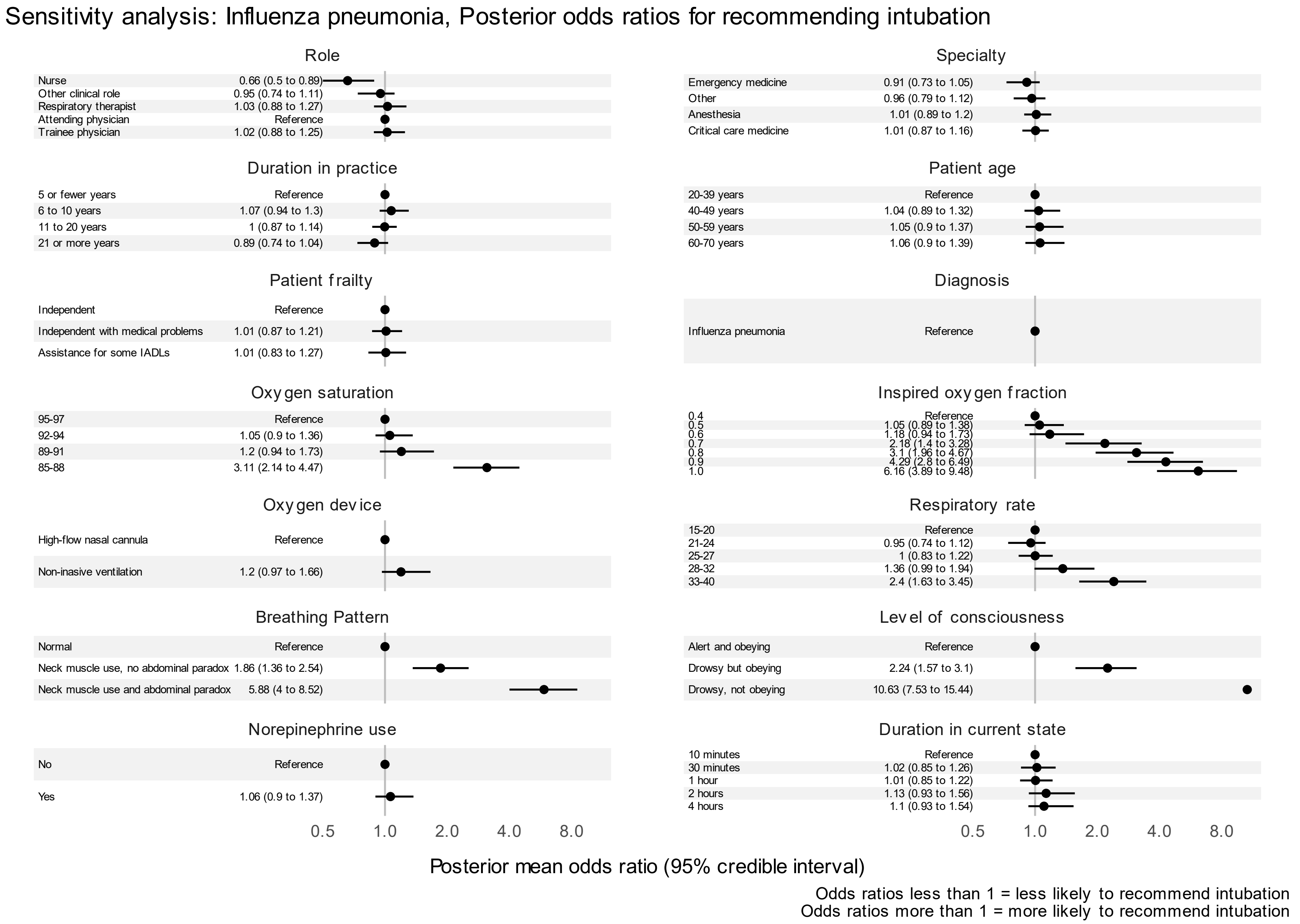

#### 11.4eFigure 19: Posterior odds ratios, COVID-19 pneumonia scenarios

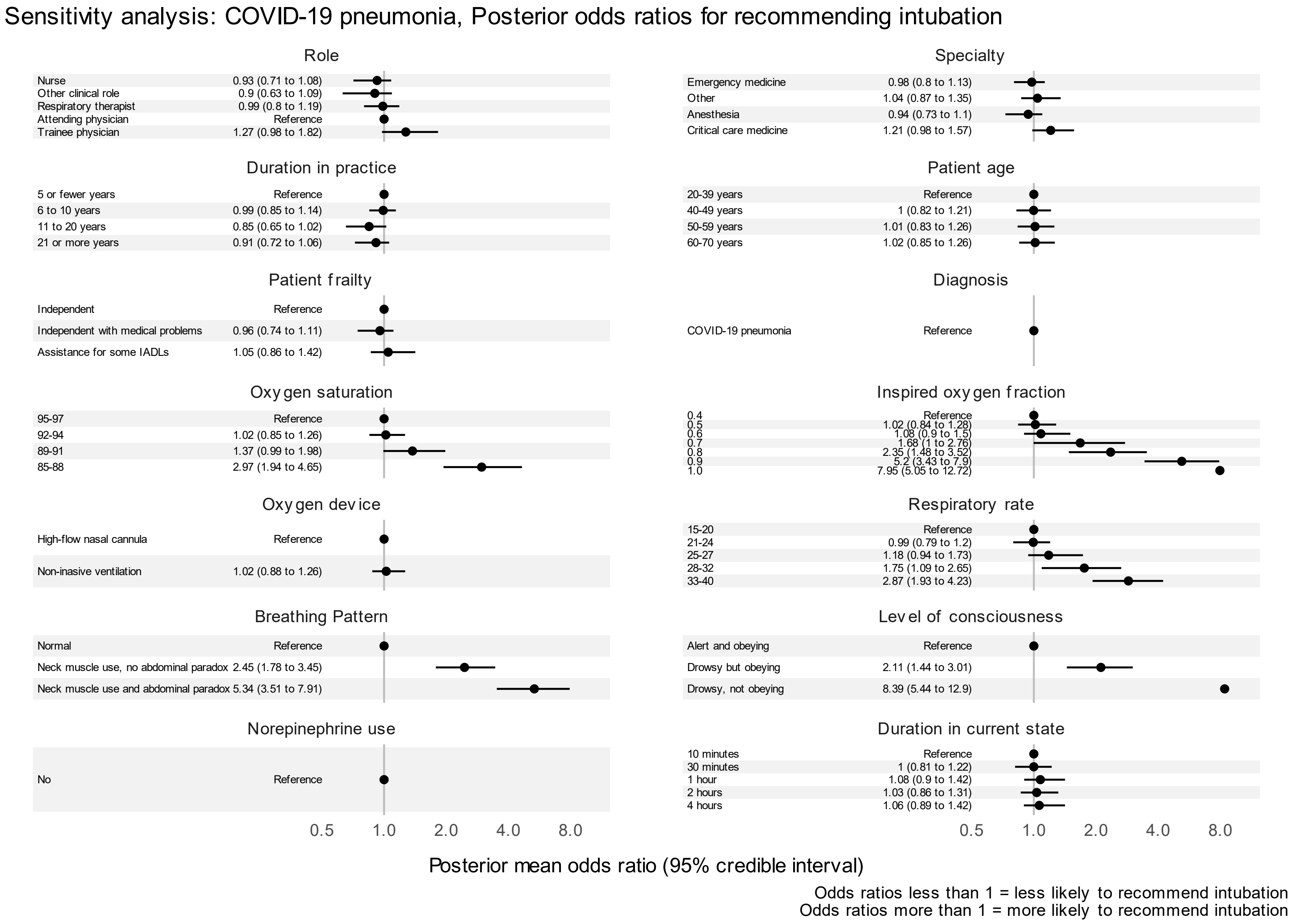

#### 11.5eFigure 20: Posterior odds ratios, pancreatitis scenarios

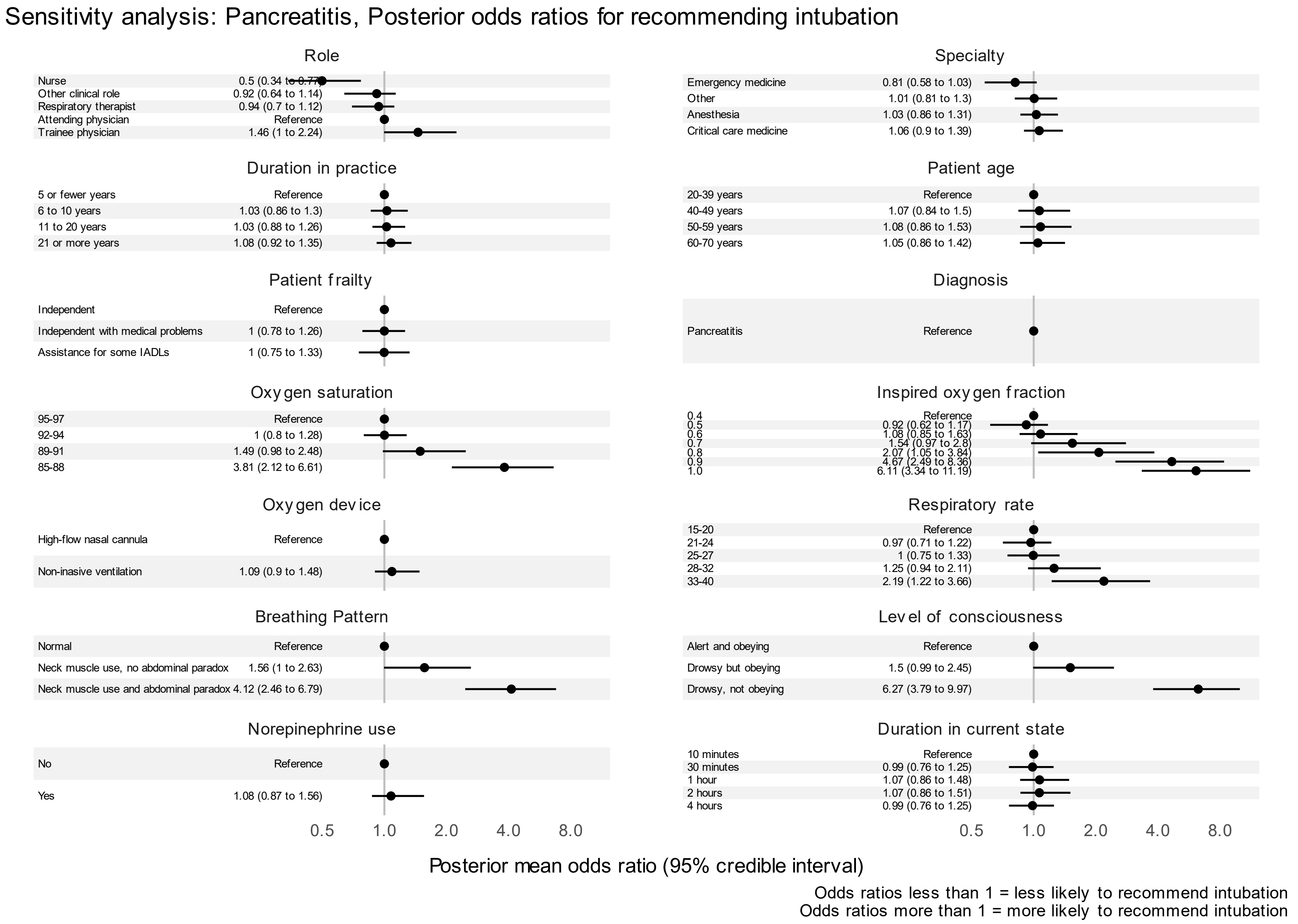

### 12 Sensitivity analysis: Assessing the proportional odds assumption

#### 12.1 Methods of testing the assumption

We performed a sensitivity analysis to assess the proportional odds assumption. The proportional odds assumption pertains to ordinal regression, or proportional odds modeling. In these circumstances, there is an ordinal outcome which can be dichotomized between any two adjacent ordinal categories to make a binary outcome. Each of these binary outcomes could be analyzed with a logistic regression model, and its fixed-effect coefficients would be odds ratios. The odds ratios from a proportional odds model can be understood as the combination of the odds ratios across all of the logistic regression models that could be formed by dichotomizing the ordinal outcome. In our case, there are four potential dichotomizations, because the ordinal scale has five elements (1 vs 2,3,4,5; 1,2 vs 3,4,5; 1,2,3 vs 4,5; 1,2,3,4 vs 5). The proportional odds assumption holds that the odds ratios for a given factor from each of those logistic regression models will be the same. This means that the change in odds of being in a higher versus lower category is the same for each level of the ordinal outcome.

To test this assumption, we fit the four logistic regression models corresponding to the dichotomizations of our ordinal outcome (Definite yes vs probable yes / uncertain / probable no / definite no; definite yes / probable yes vs uncertain / probable no / definite no; definite yes / probable yes / uncertain vs probable no / definite no; definite yes / probable yes / uncertain / probable no vs definite no). Then we inspected the corresponding odds ratios to see if they were consistent across the four logistic regression models, and similar to the findings from the primary analysis.

Of note, proportional odds models can provide accurate estimates even when there are deviations from the proportional odds assumption.(15–18) The Wilcoxon rank test is a well-known and commonly-used statistical test which depends on the proportional odds assumption, and yet its results generally hold and are valid even when that assumption is not tested.(18)

#### 12.2 Results

The proportional odds assumption was supported for all variables except respondent role of nurse. For a respondent role of nurse, the odds ratios associated with selecting “Definite yes” compared to any other option and “Definite yes” or “Probable yes” compared to any other option were both lower than 0.5, while the same odds ratios associated with selecting “Probable no” or “Definite no” compared to any other option and “Definite no” compared to any other option were very close to 1.

#### 12.3eFigure 21: Proportional odds model 1

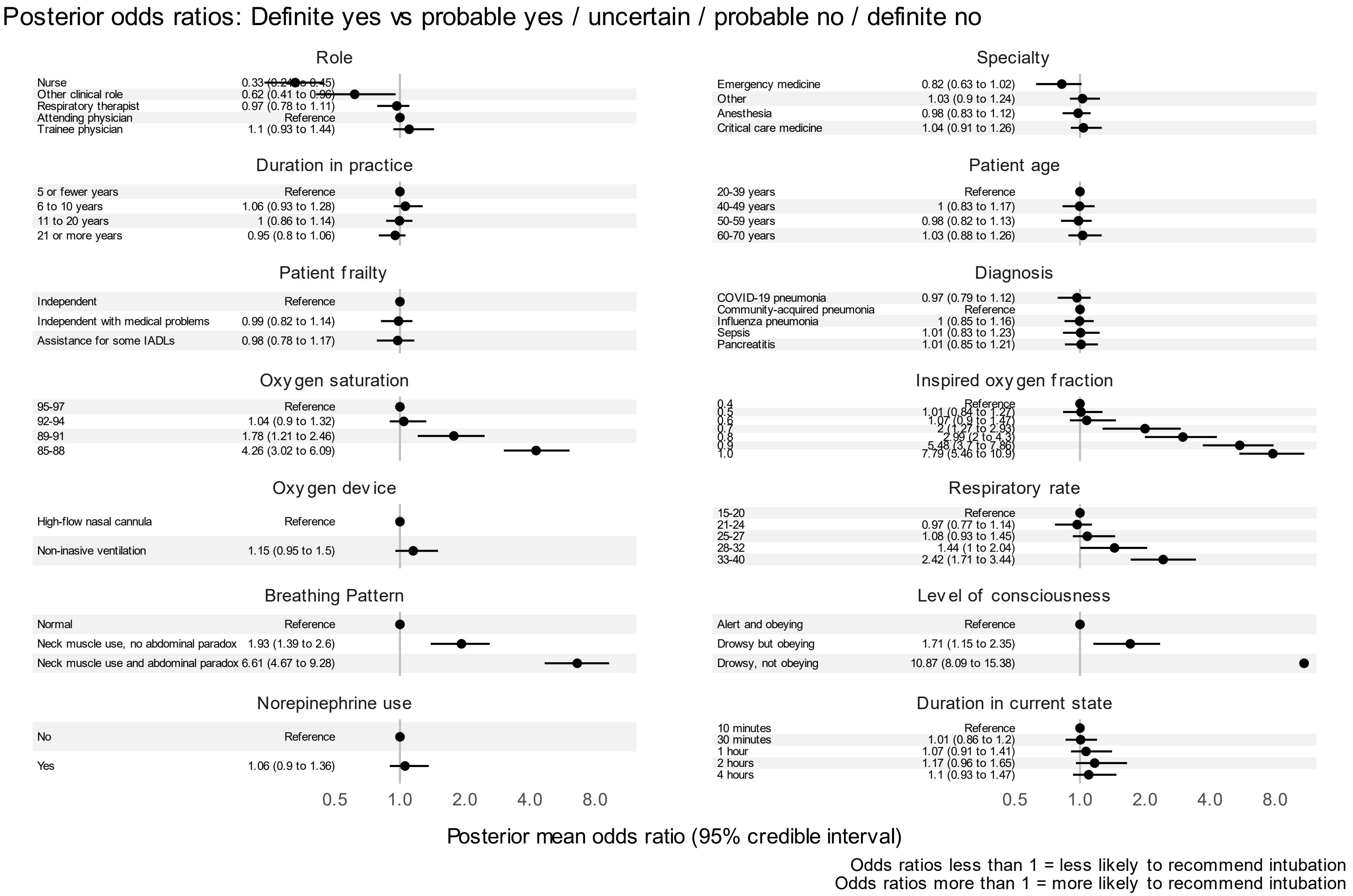

#### 12.4eFigure 22: Proportional odds model 2

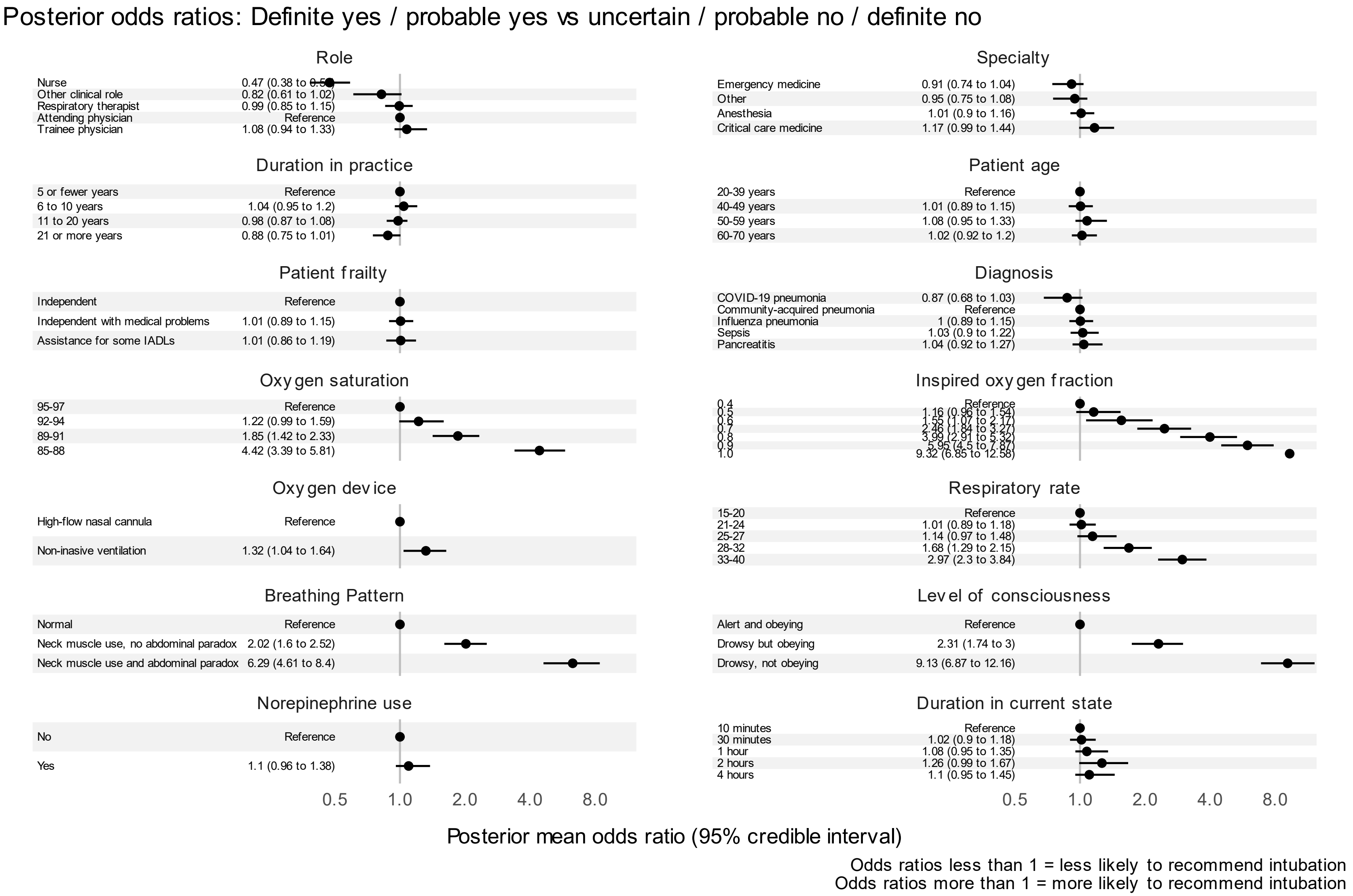

#### 12.5eFigure 23: Proportional odds model 3

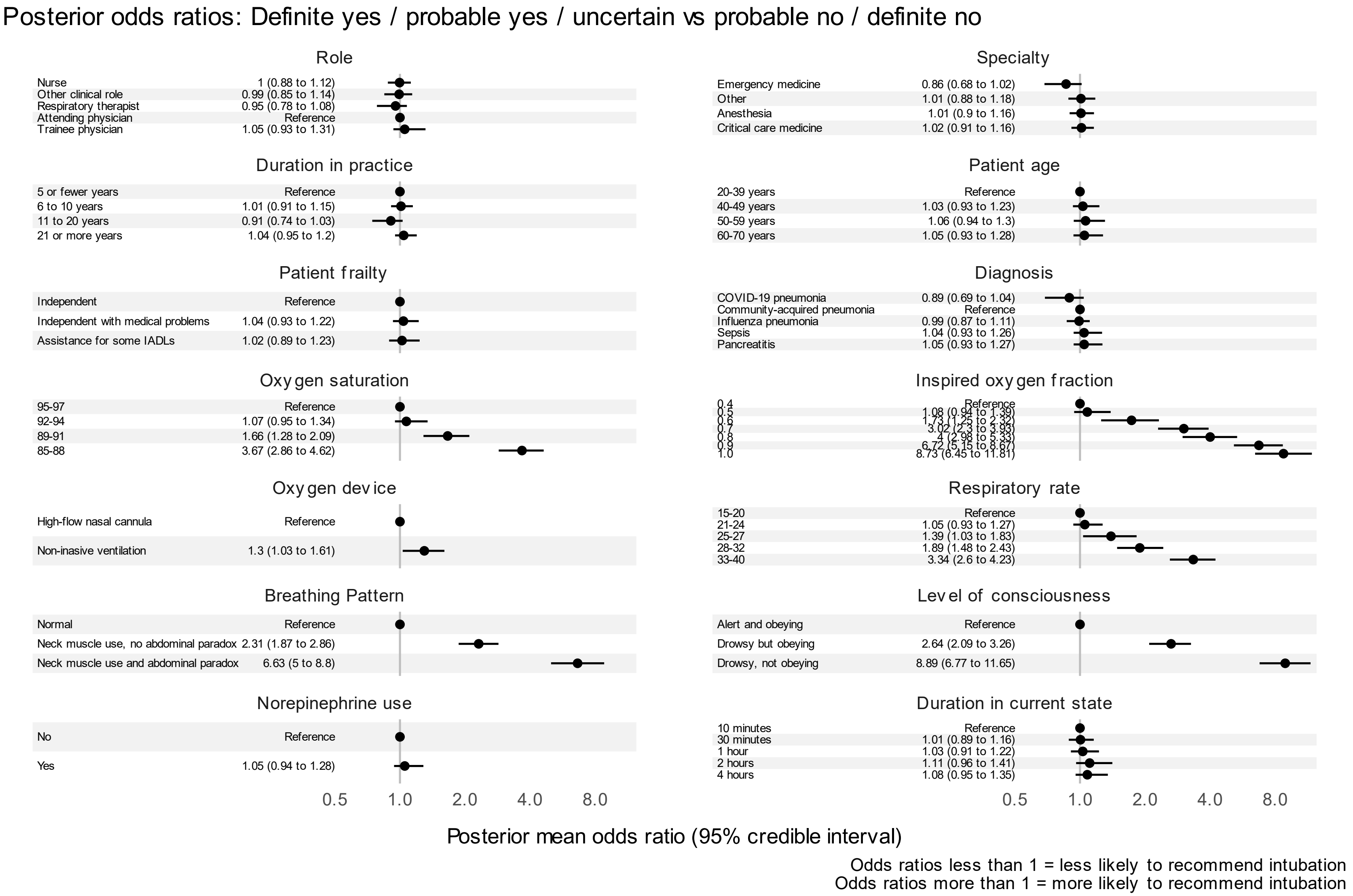

#### 12.6eFigure 24: Proportional odds model 4

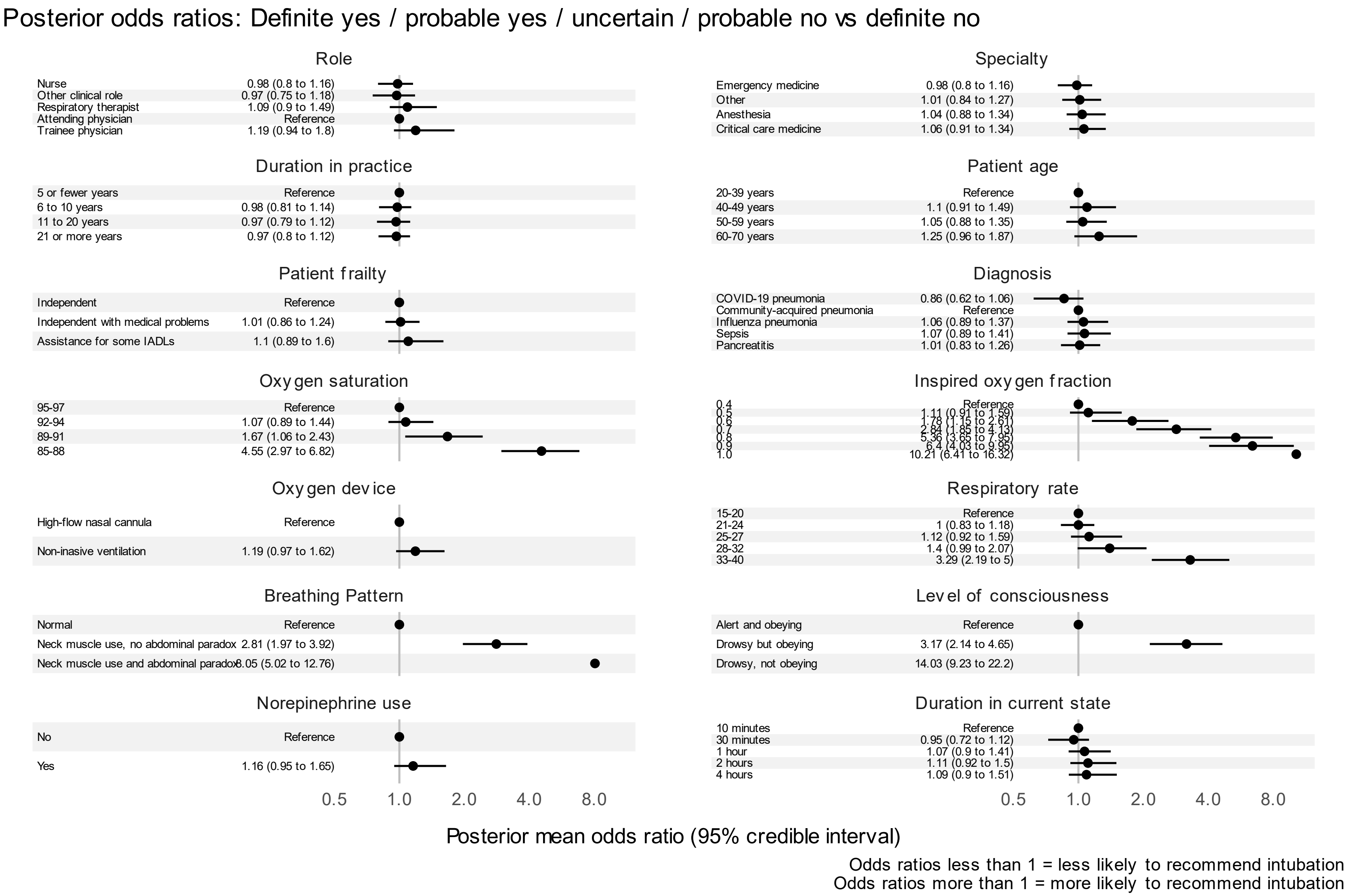
